# The Gut-Vascular Axis in Intracranial Aneurysm Rupture: A Systematic Review and Meta-analysis of Human Microbiome Evidence

**DOI:** 10.64898/2026.04.05.26350207

**Authors:** Farzan Fahim, Mahsa Hemmati, Shahriar Heshmaty, Amin Sharvirani, Amin Shahini, Ali Hosseini, Seyyed Mohammad Hosseini Marvast, Amirmahdi Mojtahedzadeh, Mohammadreza Konarizadeh, Faeze Dorisefat, Narges Maham, Abolfazl Omranisarduiyeh, Sayeh Oveisi, Farshad Fadaei Juibari, Bahador Malekipour Kashan, Guive Sharifi, Alireza Zali

## Abstract

**Background:** Intracranial aneurysm rupture is the leading cause of spontaneous subarachnoid hemorrhage and is associated with substantial mortality and long-term neurological disability. Emerging evidence suggests that the gut microbiome may influence vascular inflammation and endothelial integrity through immune and metabolic pathways, yet human evidence linking gut microbial alterations to intracranial aneurysm remains fragmented and inconsistent.

**Objective:** This systematic review and meta-analysis aimed to synthesize available human evidence on the association between gut microbiome alterations and intracranial aneurysm formation or rupture, with a primary focus on microbial dysbiosis and differences in gut microbial alpha diversity.

**Methods:** This study was conducted according to PRISMA 2020 guidelines and the protocol was prospectively registered in PROSPERO (CRD420261360785). A comprehensive search of PubMed, Scopus, Web of Science, Embase, and Cochrane CENTRAL was performed from database inception until April 1, 2026, with additional screening of grey literature sources. Observational human studies evaluating gut microbiome characteristics in patients with intracranial aneurysm were included. Mendelian randomization (MR) studies investigating genetically predicted microbial taxa and aneurysm outcomes were also reviewed. Random-effects meta-analysis using standardized mean differences (SMD) was performed for alpha diversity outcomes. MR taxa reported in at least two independent studies were quantitatively synthesized using inverse variance weighting of log-odds ratios.

**Results:** The systematic search identified 396 records. After removal of duplicates and eligibility screening, 20 studies met inclusion criteria, including 12 observational clinical studies and 8 Mendelian randomization analyses. Meta-analysis of three microbiome sequencing studies demonstrated significantly reduced gut microbial alpha diversity in patients with ruptured intracranial aneurysms compared with controls. Sensitivity analyses confirmed the robustness of pooled estimates. In addition, MR evidence identified several microbial taxa, including Ruminococcus1, Bilophila, Fusicatenibacter, and Porphyromonadaceae, as potentially protective factors against aneurysm-related outcomes. Across observational studies, gut dysbiosis was frequently associated with inflammatory pathways and alterations in microbial metabolites implicated in vascular dysfunction.

**Conclusion:** Current human evidence suggests a potential association between gut microbiome dysbiosis and intracranial aneurysm pathophysiology, particularly in relation to aneurysm rupture. Reduced microbial diversity and specific microbial taxa may influence vascular inflammation and aneurysm wall stability. However, existing evidence remains limited and heterogeneous. Large prospective cohorts and mechanistic studies are required to clarify causal relationships and evaluate whether microbiome-targeted interventions could contribute to aneurysm risk stratification or prevention strategies.

## Introduction

Intracranial aneurysms (IA) occur when a part of a cerebral artery wall weakens over time, becoming sac-shaped and bulging outward(1). In most individuals, aneurysms do not cause significant symptoms and are usually identified incidentally during brain scans for other issues; approximately 3.2% of adults worldwide have an intracranial aneurysm(2). The rupture of aneurysms is the cause of about 85% of sudden subarachnoid hemorrhages (SAH), which can lead to severe bleeding with a high risk of death, permanent brain problems, a severe decrease in quality of life, and long-term disability for individuals(3).

Some factors, such as smoking, high blood pressure, heavy alcohol consumption, plaque buildup in arteries, and a combination of genetic factors and lifestyle, are recognized today as risk factors for the development or rupture of aneurysms, given our scientific progress compared to the past. However, it is still not precisely clear why some aneurysms remain with the patient without causing any specific problem, while others suddenly rupture(4, 5).

The gut microbiome can influence how the body functions in digesting food, regulating the immune system, and controlling widespread inflammation. In some studies, this microbiome has been linked to various cerebrovascular problems, including aneurysms(6). Dietary fiber is broken down with the help of beneficial bacteria in the gut and used in the production of short-chain fatty acids (SCFA), which reduce body inflammation and also play a protective role for blood vessels. It is possible that the weakening of vessel walls and the creation of aneurysms, or even their rupture, occurs when the balance of microbes is disturbed (dysbiosis), leading to a decrease in the production of these beneficial acids and an increase in harmful byproducts(7, 8). Animal experiments conducted on this subject confirm this issue: eliminating or altering gut microbes can help reduce the likelihood of aneurysm formation and prevent their rupture(9). Some researchers have recently begun investigating whether gut microbes are involved in the progression of these aneurysms, for example, through their effects on arterial hardening or blood pressure levels(9, 10).

Many early human studies were accompanied by limitations; for example, they examined only a limited timeframe, were conducted in several limited locations worldwide, and did not fully consider factors such as individual diet, age, or ethnic differences. Despite all the advanced multi-omics work being done, there is still no firm evidence or proof that can establish a direct link between gut bacteria and whether aneurysms remain problematic or rupture (11, 12). A recent review summarized this topic, stating that the existing evidence regarding humans is still weak and we strongly need studies with longer follow-ups and actual interventional trials(13).

Ruptured aneurysms leading to subarachnoid hemorrhage result in individuals having a severely reduced chance of survival and recovery; therefore, identifying unruptured intracranial aneurysms (UIA) in the early stages and identifying external factors that are modifiable and prevent their rupture is of extraordinary importance(14). The role of the gut microbiome becomes prominent here because we can manipulate it using changes in individual diets, the consumption of probiotics, etc. Thus, it can seve as a very useful marker because we can examine it without the need for surgery or invasive procdures, or it could even become an important target in treatment. Current studies are very interesting, significant, and thought-provoking, but at the same time, they are scattered and, in some cases, not aligned or in agreement(11).

This systematic review and meta-analysis aimed to synthesize available human evidence on the relationship between gut microbiome alterations and intracranial aneurysm, with a primary focus on differences in alpha diversity and the presence of gut dysbiosis between ruptured and unruptured aneurysms. By integrating microbial diversity metrics and taxonomic findings across studies, we sought to identify microbial patterns that may act as potential risk-enhancing or protective factors for aneurysm rupture.

### Methodology

This review was conducted according to PRISMA 2020 guidelines(15). The PRISMA checklist is available in supplementary file 1. The protocol was registered in PROSPERO(16) under registration ID: CRD420261360785. The protocol registry file is available in supplementory file 2.

### Eligibility Criteria

Eligible observational studies including cohort, case-control and cross-sectional studies investigating the impact of gut microbiome alteration or dysbiosis on the risk of intracranial aneurysm formation or rupture were included. These studies enrolled human subjects with diagnosed intracranial aneurysms (e.g. CT, MRI, angiogram) assessed regarding their gut microbiome composition or related metabolites using valid techniques (e.g. 16S rRNA sequencing, shotgun metagenomics, or metabolomic analyses). All studies from inception until April 1, 2026 were included. All languages were included and No restrictions were applied. Animal studies, case reports, case-series with fewer than 10 patients, conference papers, reviews, editorials and letters were excluded. Studies that did not investigate gut microbiome characteristics with standard techniques, did not include patients with ICA with standard diagnosis or did not link the gut microbiome to ICA outcomes were excluded.

### Search Strategy

A comprehensive search in major databases including PubMed, Scopus, Web of Science, Embase and Cochrane CENTRAL was done on April 1, 2026. Grey literature including bioRxiv, medRxiv and ClinicalTrial.gov were also searched for preprints and ongoing studies but they were not included in the final data synthesis since they have not been peer-reviewed. References of included studies were also checked by two reviewers as an additional source.

Our search core consisted of “gut microbiome”, “intracranial aneurysm”,” formation” or “rupture” in variations. PubMed search term is presented below and you can access search terms for other databases in supplementary 3.

PubMed search term: ( Gut Microbiome[tiab] OR Gut Microbiota[tiab] OR Gut flora[tiab] OR Gut microflora[tiab] OR gut dysbiosis[tiab] OR intestinal microbiota[tiab] OR intestinal microbiome[tiab] OR intestinal flora[tiab] OR intestinal microflora[tiab] OR intestinal dysbiosis[tiab] OR gastrointestinal microbiome[tiab] OR gastrointestinal microbiota[tiab] OR gastrointestinal flora[tiab] OR gastrointestinal microflora[tiab] OR gastrointestinal dysbiosis[tiab] OR gastric microbiome[tiab] OR gastric microbiota[tiab] OR gastric flora[tiab] OR gastric microflora[tiab] OR gastric dysbiosis[tiab] OR enteric microbiome[tiab] OR enteric microbiota[tiab] OR enteric flora[tiab] OR enteric microflora[tiab] OR enteric dysbiosis[tiab] OR Microflora[tiab] OR Microbiota[tiab] OR Microbiome[tiab] OR flora[tiab] OR Dysbiosis[tiab] OR microbiota[tiab] OR microbiome[tiab] OR microflora[tiab] OR flora[tiab] OR gut-brain axis[tiab] OR "Microbiota"[Mesh] OR "Gastrointestinal Microbiome"[Mesh] OR "brain-gut axis"[Mesh] OR "Dysbiosis"[Mesh] ) AND ( "Intracranial Aneurysm*"[tiab] OR "Cerebral Aneurysm*"[tiab] OR "Brain Aneurysm*"[tiab] OR intracranial mycotic aneurysm*[tiab] OR anterior cerebral artery aneurysm*[tiab] OR ACA aneurysm*[tiab] OR posterior cerebral artery aneurysm*[tiab] OR PCA aneurysm*[tiab] OR anterior communicating artery aneurysm*[tiab] OR basilar artery aneurysm*[tiab] OR middle cerebral artery aneurysm*[tiab] OR MCA aneurysm*[tiab] OR posterior communicating artery aneurysm*[tiab] OR berry aneurysm*[tiab] OR subarachnoid hemorrhage[tiab] OR SAH[tiab] OR "Intracranial Aneurysm"[Mesh] OR "Subarachnoid Hemorrhage"[Mesh] ) AND ( formation[tiab] OR rupture[tiab] OR ruptured[tiab] OR growth[tiab] OR risk[tiab] OR association[tiab] OR prognosis[tiab] )

### Selection Process

After importing all the search results into Rayyan, the deduplication process was done manually. Two reviewers (MRK and AH) independently screened titles and abstracts based on predefined eligibility criteria. All excluded studies in this stage were documented in a title abstract exclusion sheet provided by senior author (FF) with exclusion reason. Any conflicts were resolved via discussion and a third person opinion (MH). These sheets are presented in supplementary file 4. Full texts of the remaining studies were then screened by two independent reviewers (ASharvirani and AShahini). All the studies in this process were documented in full text exclusion sheet provided by senior author (FF) (including title, author, doi, origin, exclusion reason) and inclusion sheet (including title, author, doi, origin, population, exposure,outcome, comparison group). These sheets are presented in supplementary files 5,6. All the conflicts were resolved by a third person opinion (MH) and final included studies are presented in supplementary file 5.

### Data Collection

Two reviewers (ASharvirani and ShH) independently extracted data according to predefined extraction sheet provided by senior author (FF) and it was reviewed by a third person (FF). All results compatible with each outcome domain were sought and if multiple measures were reported the most adjusted model was extracted. In case of missing or unclear data authors were contacted via email.

### Data Items

Data were sought in 9 domains including study characteristics, population characteristics, aneurysm characteristics, gut microbiome characteristics, outcomes, biological mechanisms, interventions, effect size and confounding factors. Study characteristics included title, author, publication date, DOI, ethical approval, country, study design, center type, time period of data collection. In population characteristics inclusion and exclusion criteria, follow up duration, number of patients, mean age, sex, BMI, hypertension, smoking, diabetes, dyslipidemia, recent antibiotics, drugs (PPIs, statins, antihypertensive drugs) were sought for each group including cases (ruptured aneurysms and unruptured aneurysms) and controls individually if available. Aneurysm characteristics included size, location, shape, number (multiple or singular). For gut microbiota sampling time, sampling type, sequencing method, alpha diversity, Firmicutes abundance, Bacteriodetes abundance, F/B ratio, presence of dysbiosis were extracted. In terms of outcomes vasospasm incidence, delayed stroke, hydrocephalus incidence, rebleeding incidence, mortality at 3 time points of hospital, 3 months and 6 months, ICU stay, hospital stay, MRS<2 at discharge, Hunt-Hess grade, Fishers Grade, Aneurysm treatment was considered. Biologic mechanisms included mechanism category, signaling pathway, biomarkers, direction of change, significance, metabolite type, measurement and relationship. For interventions we extracted types of intervention if presented. For effect size number of dysbiosis with upper and lower confidence interval, microbiome toxonomy with upper and lower CI, alpha diversity, effect size, inflammatory biomarkers and metabolites were extracted. Finally, to assess confounders risk of bias, adjusted confounding factors, adjustment method, funding source, conflict of interest and limitations were sought. Each item was sought individually for 3 groups including ruptured and unruptured aneurysms as case groups and control group if applied. These extraction sheets are available in supplementary file 7.

### Risk Of Bias

Risk of bias was accessed by two independent reviewers (NB and NM) using Cochrane Quality in Prognosis Studies tool (QUIPS). This tool assesses the risk of bias in six domain including study participation, study attrition, prognostic factor measurement, outcome measurement, study confounding, statistical analysis and reporting. Each domain consists of questions to evaluate risk of bias and the answers to these questions are available on supplementary files 8.

### Data Synthesis and Meta-Analysis

#### Alpha Diversity

A quantitative meta-analysis was conducted by RevMan4.5.1 application to evaluate differences in gut microbiome alpha diversity between patients with ruptured intracranial aneurysm and control groups across published microbiome sequencing studies. Eligible studies were identified that reported alpha diversity indices comparing aneurysm rupture cases with controls. Three independent studies were included in the final synthesis: Xu et al. (2024)(17), Csecsei et al. (2025)(12), and Zhang et al. (2025)(18). For studies reporting alpha diversity values as mean ± standard deviation, these values were directly extracted. However, in studies where alpha diversity was reported only graphically (e.g., boxplots), numerical values were digitized from the figures using WebPlotDigitizer software. Extracted statistics included the median, interquartile range, and whisker values when available. These values were subsequently converted to estimated mean and standard deviation using established statistical methods described by Wan et al. (2014)(19) and Luo et al. (2018)(20). This approach allows approximate reconstruction of continuous summary statistics when raw data are not reported. Because different alpha diversity metrics and scales were used across studies, the standardized mean difference (SMD) was calculated as the pooled effect size. Meta-analysis was performed using the inverse-variance method. Between-study heterogeneity was assessed using Cochran’s Q test and quantified using the I² statistic and τ². Given expected methodological variability between microbiome studies (including sequencing platforms, diversity indices, and study populations), a random-effects model was applied for the primary analysis. To evaluate the robustness of the pooled estimate, sensitivity analysis was performed using a leave-one-out approach, in which the meta-analysis was repeated after sequential exclusion of each individual study. Influence diagnostics were conducted to identify potentially influential or outlying studies using multiple metrics including studentized residuals, Cook’s distance, DFFITS, covariance ratios, and leverage values. Potential publication bias was visually assessed using funnel plot inspection, plotting standardized mean differences against their standard errors.

#### Mendelian Randomization Taxa Meta-Analysis

To investigate potential causal relationships between gut microbial taxa and aneurysm-related outcomes, results from Mendelian randomization (MR) studies were synthesized. MR studies reporting associations between genetically predicted gut microbiota abundance and aneurysm outcomes were reviewed. Taxa were included in the quantitative synthesis only if they were reported in at least two independent MR studies with comparable taxonomic classification. Four microbial taxa fulfilled this criterion: Ruminococcus1, Bilophila, Fusicatenibacter, and Porphyromonadaceae. For each study, effect estimates were extracted as odds ratios (OR) with corresponding 95% confidence intervals derived from inverse-variance weighted MR analyses.Meta-analysis was performed using inverse-variance weighting on the logarithmic scale of the odds ratios. Random-effects models were applied to account for potential differences in genetic instruments and study populations.Between-study heterogeneity was assessed using Cochran’s Q statistic and the I² index.

## RESULT

The systematic literature search identified 396 records from multiple databases and registries. After removing 173 duplicates, 223 records were screened by title and abstract, resulting in the exclusion of 188 studies. Full-text articles were retrieved for 35 records; one could not be obtained. Following eligibility assessment of 34 studies, 14 were excluded (9 lacked aneurysm-related outcomes, and 5 lacked microbiome assessments). Ultimately, 20 studies met the inclusion criteria and were included in the qualitative and quantitative synthesis: 12 observational clinical studies and 8 two-sample Mendelian randomization (MR) analyses. The study selection process is illustrated in Figure 1 (PRISMA flow diagram).the full version of result is provided in supplementory file 9.

**Figure 1.**
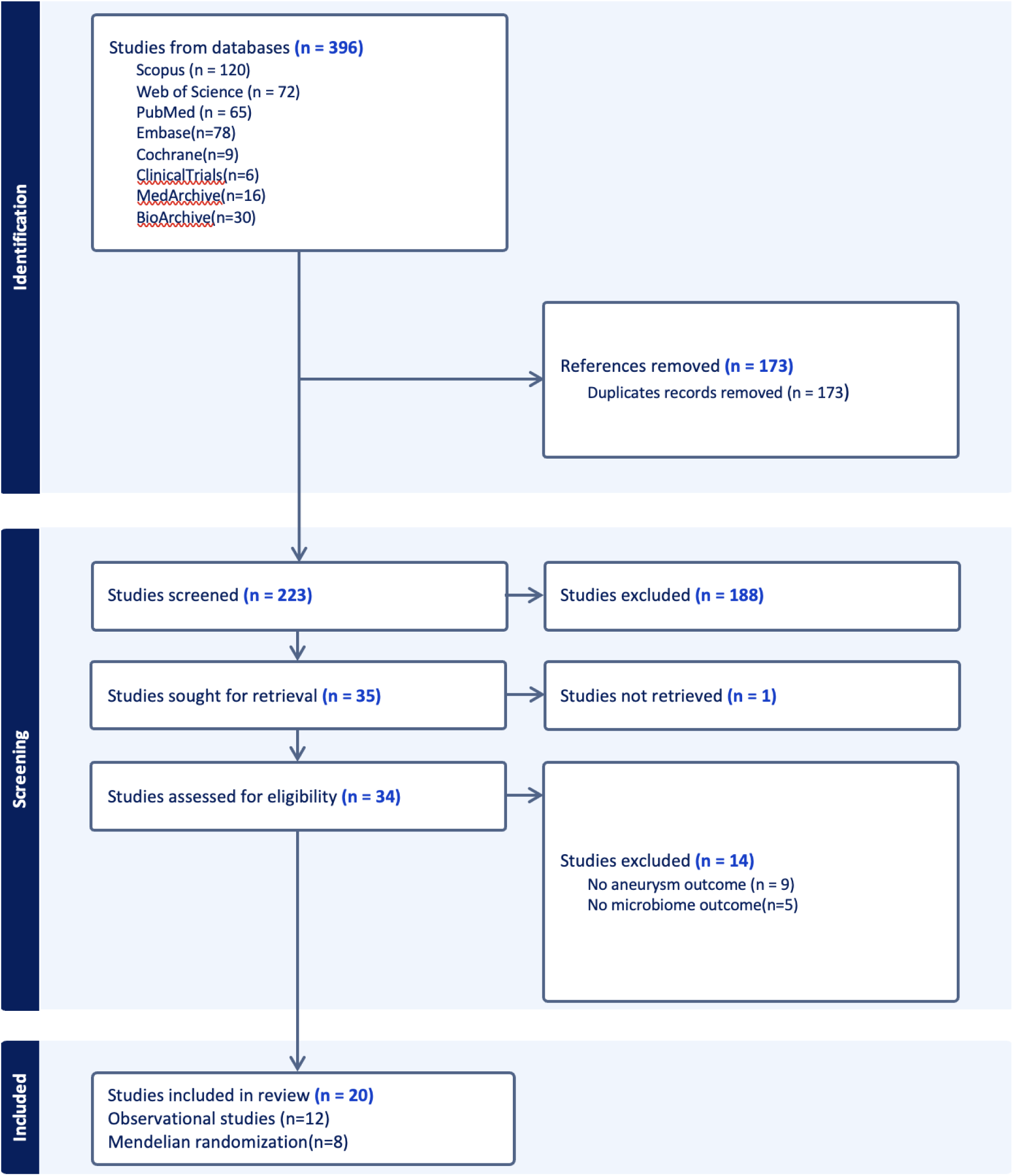
PRISMA flowchart

### Study characteristics

Key characteristics of the included studies are summarized in Table 1. The 12 clinical and experimental studies were predominantly observational cohort or case–control investigations conducted in China, Japan, Hungary, and Brazil. These studies examined gut microbiota profiles in several intracranial aneurysm (IA), related contexts, including unruptured intracranial aneurysm (UIA) versus healthy controls (Li 2020 (7); Du 2024 (21)), ruptured versus unruptured IA (Csecsei 2025 (12); Kawabata 2022(11)), aneurysmal subarachnoid hemorrhage (aSAH) (Zhang 2025(18)), IA in patients with autosomal dominant polycystic kidney disease (ADPKD) (Fukuda 2025(22)), and the presence of bacterial DNA within aneurysm wall tissue (Rabelo 2025(23)). Sample sizes across these studies ranged from 21 to 280 participants. Gut microbiota composition was most commonly assessed using 16S rRNA gene sequencing, while shotgun metagenomic sequencing was performed in at least one study (Li 2020(7)), and targeted metabolomics was incorporated in studies by Du (21)(2024) and Zhang (2025)(18).

**Table 1.**
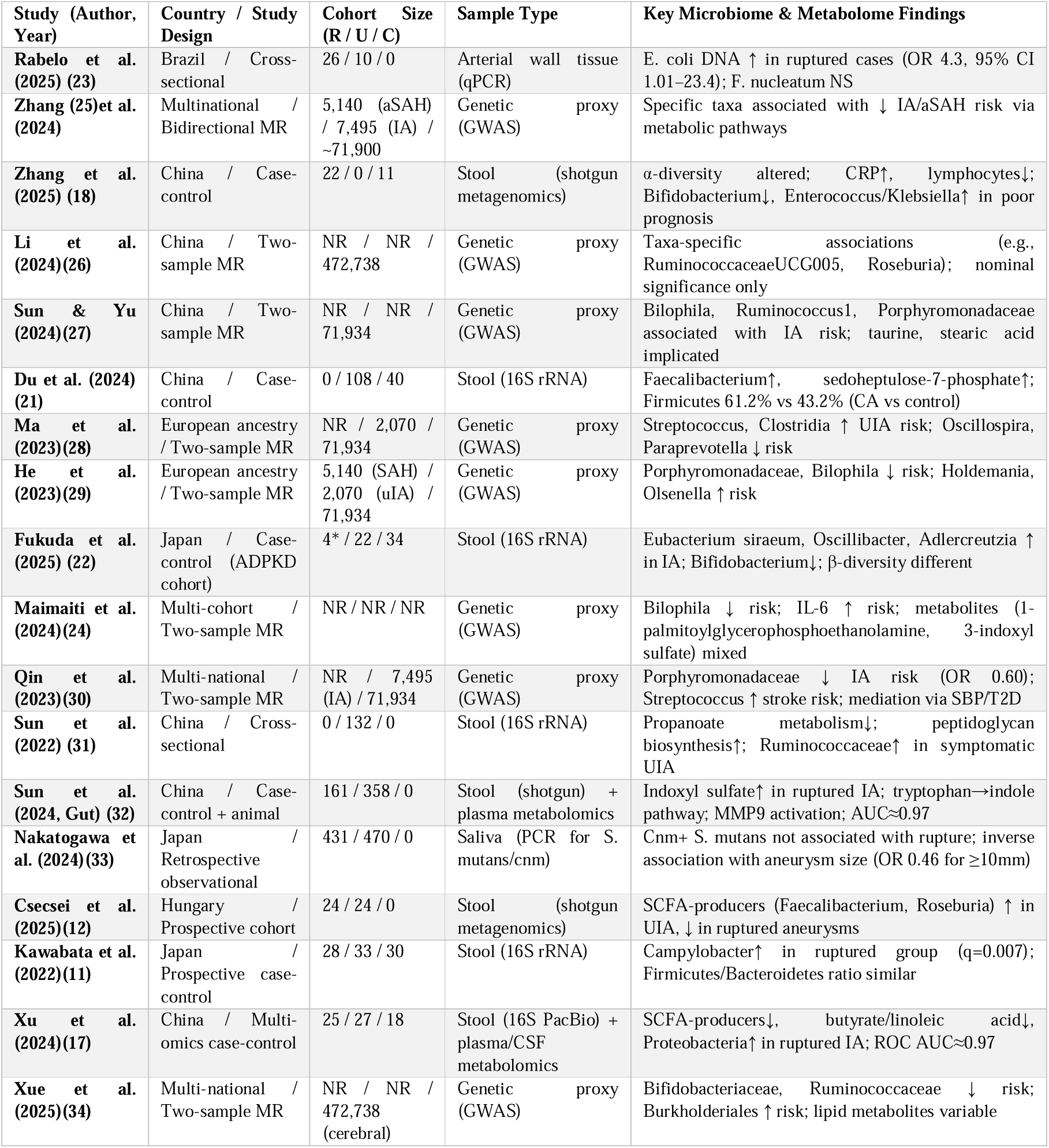
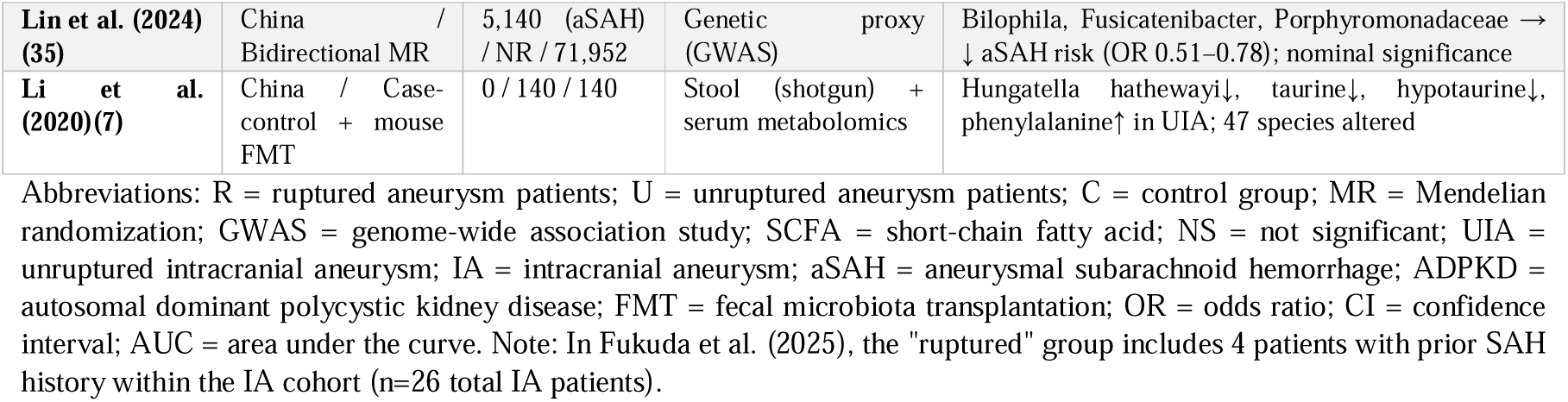
Summary of Studies on Gut Microbiota and Intracranial Aneurysms.

The 8 Mendelian randomization studies uniformly applied a two-sample MR framework. Exposures were defined as genetically predicted gut microbiota traits derived from the MiBioGen consortium, which includes approximately 5,000 individuals, while aneurysm-related outcomes were based on large Genome-Wide Association Studies (GWAS) of IA and/or aSAH. These outcome GWAS predominantly included European-ancestry participants, with case numbers typically ranging from approximately 2,000 to 5,000 and control sample sizes exceeding 70,000. One large-scale MR study (Maimaiti 2024(24)) included up to 306,882 individuals. All MR analyses were conducted in European populations, which has implications for the generalizability of the findings.

### Patient Characteristics

Clinical demographic and baseline data were available from the 12 observational studies, encompassing approximately 1,580 patients with intracranial aneurysms (IA) and 450 control participants. The eight Mendelian randomization studies utilized summary-level genome-wide association data and therefore did not contribute individual-level clinical characteristics.

### Demographics and Anthropometrics

Across clinical cohorts reporting age data (n = 7 studies), the mean age ranged from 55.8 ± 12.1 years (symptomatic UIA in Sun et al. 2022(31)) to 69.1 ± 9.9 years (unruptured IA in Kawabata et al. 2022(11)), with most cohorts clustering in the late 50s to mid-60s. Sex distribution was reported in 8 studies: male proportions ranged from 21.7% (symptomatic UIA, Sun et al. 2022(31)) to 48% (unruptured IA, Xu et al. 2024(17)) in aneurysm cohorts, and from 28% to 45.5% in control groups were reported. Body mass index was documented in 4 studies, with mean values ranging from 21.9 ± 4.0 kg/m² (ruptured IA, Kawabata et al. 2022(11)) to 22.99 ± 1.48 kg/m² (aSAH, Zhang et al. 2025(18)), indicating a generally normal-weight population.

### Comorbidities and Vascular Risk Factors

Hypertension was the most consistently reported comorbidity, with prevalence ranging from 42.4% (unruptured IA, Kawabata et al. 2022(11)) to 83% (unruptured IA, Csecsei et al. 2025(12)) across studies; notably, hypertension was more prevalent in cohorts enriched for ruptured aneurysms or acute presentations. Smoking history was reported in 7 studies, ranging from 7.0% (asymptomatic UIA, Sun et al. 2022(31)) to 58% (ruptured IA, Csecsei et al. 2025(12)). Diabetes mellitus prevalence varied from 0% (controls in Zhang et al. 2025(18) and Xu et al. 2024(17), by exclusion criteria) to 19% (unruptured IA, Xu et al. 2024(17)). Dyslipidemia was reported in 5 studies, with prevalence between 3.5% (asymptomatic UIA, Sun et al. 2022(31)) and 33.8% (unruptured IA, Nakatogawa et al. 2024(33)).

### Aneurysm Morphology and Clinical Presentation

Aneurysm size data were available from 6 studies. Ruptured aneurysms were consistently larger than unruptured lesions: mean diameters ranged from 5.8 ± 3.2 mm (Kawabata et al. 2022(11)) to 8.03 ± 4.43 mm (Xu et al. 2024(17)) in ruptured cohorts versus 3.7 ± 1.6 mm (Kawabata et al. 2022(11)) to 6.0 ± 3.8 mm (Sun et al. 2022(31)) in unruptured cohorts. The vast majority of aneurysms were located in the anterior circulation (75–96.4% across reporting studies), most frequently involving the internal carotid artery, middle cerebral artery, and anterior/posterior communicating arteries. Multiple aneurysms were identified in 12–30% of patients across studies reporting this variable. Morphologically, irregular contours or daughter sac formation were more frequently reported in ruptured cases (e.g., 59.6% vs. 31.3% in Nakatogawa et al. 2024(33)), consistent with established biomechanical risk profiles for rupture.

### Data Completeness & Reporting Patterns

Demographic and clinical data were incompletely reported across studies. Age was missing in 4/11 observational studies; BMI was reported in only 4/11; and medication histories (antibiotics, statins, antihypertensives) were inconsistently documented. Notably, several studies excluded patients with recent antibiotic use (<1 month), which may limit generalizability to broader clinical populations. Despite these gaps, the available data indicate a clinically coherent population characterized by advanced age, high prevalence of hypertension and smoking, and anterior-circulation-predominant aneurysm morphology, aligning with established epidemiological patterns for intracranial aneurysm disease.

### Gut microbiota composition and diversity in intracranial aneurysm

Across the available clinical and experimental studies, patients with intracranial aneurysms demonstrated gut microbiome dysbiosis compared with healthy controls, although specific taxa and diversity indices varied by cohort. In UIA populations, Li et al. (2020) (7) reported enrichment of pro-inflammatory genera such as Escherichia–Shigella and Enterococcus, alongside depletion of SCFA-producing taxa including Faecalibacterium and Hungatella hathewayi. Their fecal microbiota transplantation experiments showed that mice receiving microbiota from UIA patients developed significantly more severe aneurysms than controls (p < 0.05), supporting a causal contribution of dysbiosis. Du et al. (2024) (21) confirmed similar dysbiotic patterns in UIA and additionally identified a distinct serum metabolomic signal, particularly involving sedoheptulose-7-phosphate, suggesting microbiota-modulated metabolic alterations in aneurysm biology. Differences according to rupture status were demonstrated in two independent studies. Kawabata et al. (2022) (11) found dysbiosis exclusively among patients with ruptured IA, who also had significantly larger aneurysms (5.8 ± 3.2 mm vs. 3.7 ± 1.6 mm; p < 0.05). Csecsei et al. (2025) (12) likewise reported distinct microbial community structures in ruptured versus unruptured IA, with rupture associated with a marked reduction in butyrate-producing bacteria, suggesting loss of SCFA-mediated vascular protection. In patients with aSAH, Zhang et al. (2025) (18) identified pronounced overgrowth of Escherichia–Shigella, depletion of SCFA-producing taxa, reduced fecal butyrate and propionate, and elevated LPS-related metabolites, all correlating with systemic inflammation. Gut microbiota differences were also observed in high-risk ADPKD patients. Fukuda et al. (2025) (22) found significant compositional differences between ADPKD patients with and without IA, despite similar vascular risk profiles, suggesting a potential microbiome-related contribution to aneurysm formation independent of traditional factors. Supporting a possible local role for bacteria, Rabelo et al. (2025) (23) detected Escherichia coli and Fusobacterium nucleatum DNA within aneurysm wall tissue; confidence intervals for E. coli (1.01–23.4) were compatible with a positive association, suggesting bacterial components may contribute to local inflammatory injury, though causality could not be established. Several studies described changes in the Firmicutes/Bacteroidetes ratio and alpha-diversity; however, findings were inconsistent and quantitative data incomplete, preventing formal meta-analysis of these parameters.

### Causal inference from Mendelian randomization

A total of 8 Mendelian randomization (MR) studies evaluated the potential causal relationship between gut microbiota and the risk of intracranial aneurysm (IA) and/or aneurysmal subarachnoid hemorrhage (aSAH) using genetic instruments for microbial traits. Most studies reported statistically significant associations between specific microbial taxa and aneurysm outcomes, generally based on inverse-variance–weighted MR analyses with supportive sensitivity tests. However, detailed numerical effect estimates were not consistently available, limiting comprehensive quantitative synthesis. The clearest quantifiable finding was reported by Qin et al. (2023) (30), who showed that a higher genetically predicted abundance of the bacterial family Porphyromonadaceae was associated with a reduced risk of IA (95% CI 0.44–0.83), suggesting a protective causal effect. Overall, MR evidence supports a potential causal role of gut microbiota in aneurysm pathogenesis, with proposed mechanisms including modulation of inflammatory pathways (TLR4, NF-κB, NLRP3 inflammasome), SCFA-mediated endothelial protection, TMAO-related oxidative stress, and microbiota-dependent regulation of blood pressure and vascular tone.

### Alpha diversity

Three observational studies comprising a total of 202 participants were included in the meta-analysis. Across these studies, alpha diversity indices were compared between aneurysm rupture cases and control groups. The pooled analysis demonstrated a reduction in alpha diversity among patients with aneurysm rupture compared with controls. The overall pooled standardized mean difference was −0.94 (95% CI −1.31 to −0.57, p < 0.0001), indicating a large effect size and may suggesting substantially reduced microbial diversity in the rupture group. Study specific effect sizes were as follows: Xu et al. (2024)(17) reported SMD −0.96 (95% CI −2.06 to 0.14), Csecsei et al. (2025)(12) reported SMD −1.31 (95% CI −2.76 to 0.85), and Zhang et al. (2025)(18) reported SMD −0.50 (95% CI −1.93 to −0.70). Moderate heterogeneity was observed across studies (I² = 48.4%, τ² = 0.1045, p = 0.144). This suggests moderate between study variability, likely reflecting differences in study populations, sequencing methods, diversity metrics, and timing of sample collection relative to aneurysm rupture (fig.3). Sensitivity analysis using the leave one out method (fig.5) demonstrated that the pooled effect remained statistically significant regardless of which study was removed. When Xu et al. (2024)(17) was excluded, the pooled estimate was SMD −0.72 (95% CI −1.19 to −0.26; p = 0.0022). Exclusion of Csecsei et al. (2025)(12) yielded a pooled estimate of SMD −1.24 (95% CI −1.73 to −0.76; p < 0.0001). Excluding Zhang et al. (2025)(18) resulted in a pooled estimate of SMD −0.88 (95% CI −1.30 to −0.46; p < 0.0001). These results confirm the robustness of the observed association. Influence diagnostics (fig.6) showed no evidence that any single study exerted disproportionate influence on the pooled estimate. Studentized residuals, Cook’s distance, and leverage values were all within acceptable ranges. Visual inspection of the funnel plot (fig.4) did not reveal clear evidence of publication bias. However, interpretation of funnel plot symmetry is limited due to the small number of included studies. This meta-analysis demonstrates a consistent and substantial reduction in gut microbiome alpha diversity among patients with aneurysm rupture compared with control groups. The pooled standardized mean difference of −0.94 indicates a large effect size, may suggest that microbial diversity may decreased in the rupture population.

**Figure 2.**
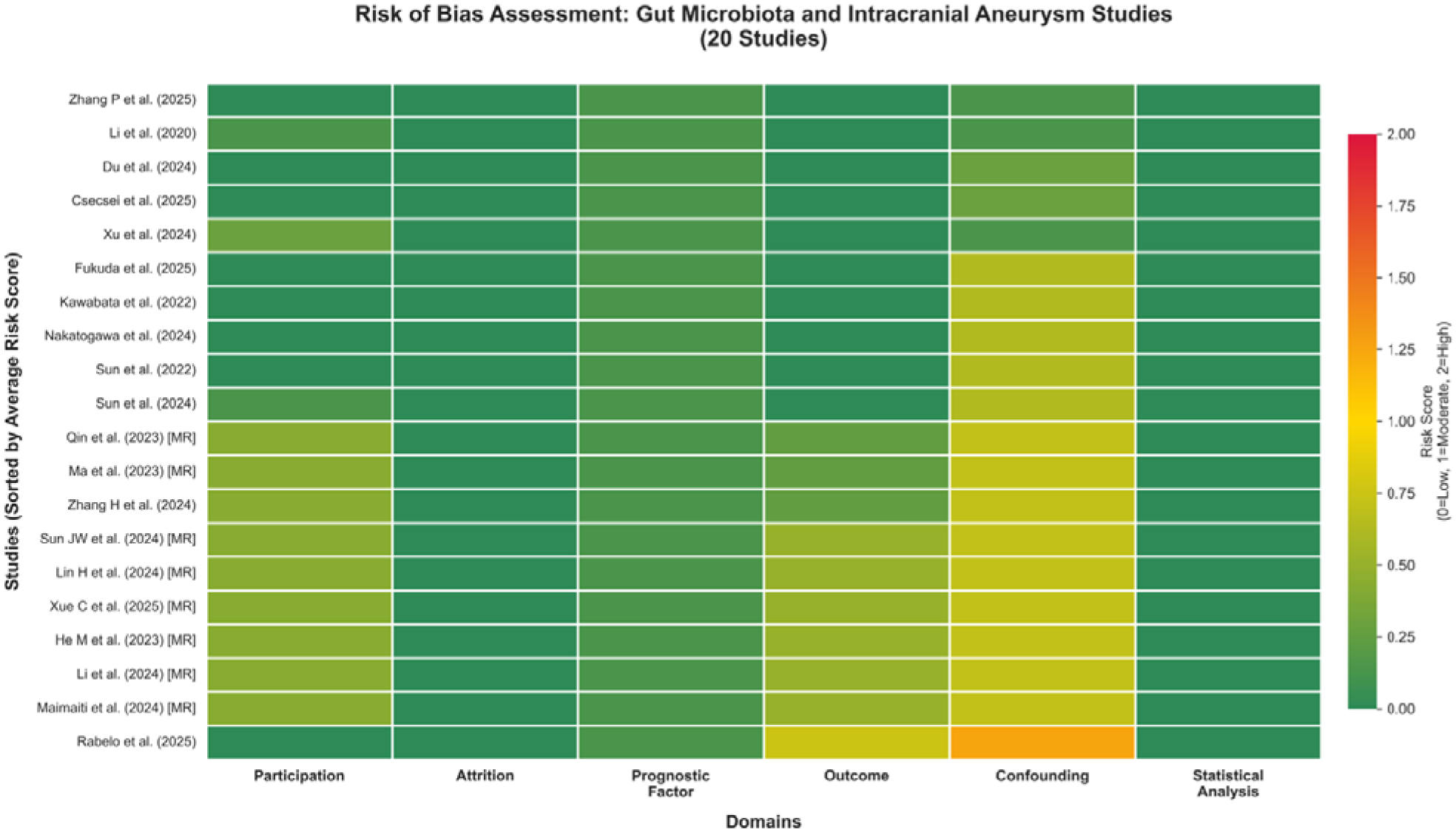
Risk of bias assessment of included studies. Each cell represents the risk score for one of six domains: Study Participation, Study Attrition, Prognostic Factor Measurement, Outcome Measurement, Study Confounding, and Statistical Analysis. Color coding: Green = Low risk (score 0.0–0.5); Yellow = Moderate risk (0.5–1.5); Red = High risk (1.5–2.0). Note: Confounding was the most frequently problematic domain, particularly regarding unmeasured microbiome-specific confounders (diet, medications, oral health). Nakatogawa et al. (2024) assessed oral (not gut) microbiota; moderate risk reflects unmeasured oral hygiene/periodontal confounders.

**Figure 3.**
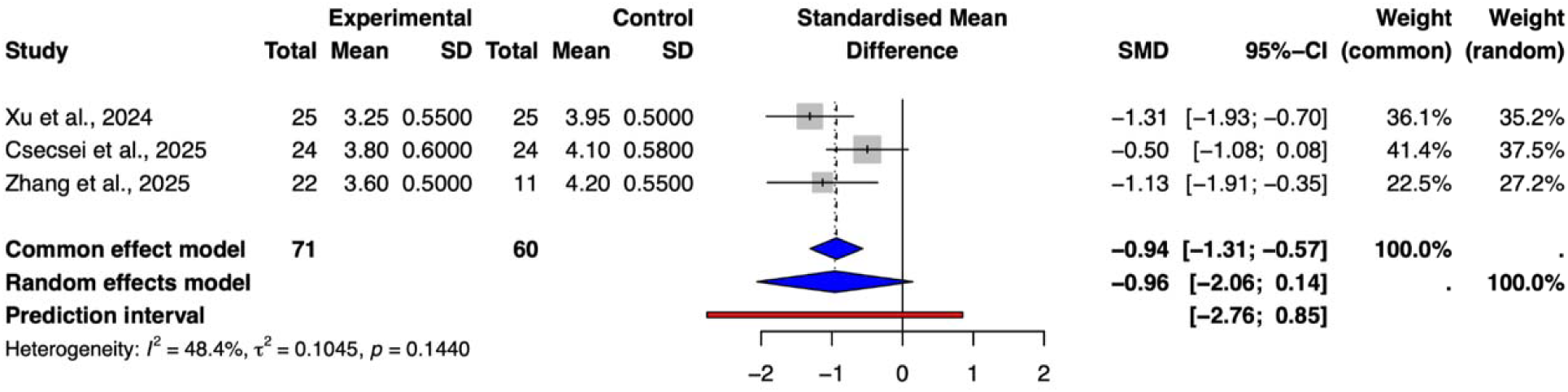
Forest plot illustrating the pooled standardized mean difference in gut microbiome alpha diversity between patients with ruptured intracranial aneurysms and control groups.

**Figure 4.**
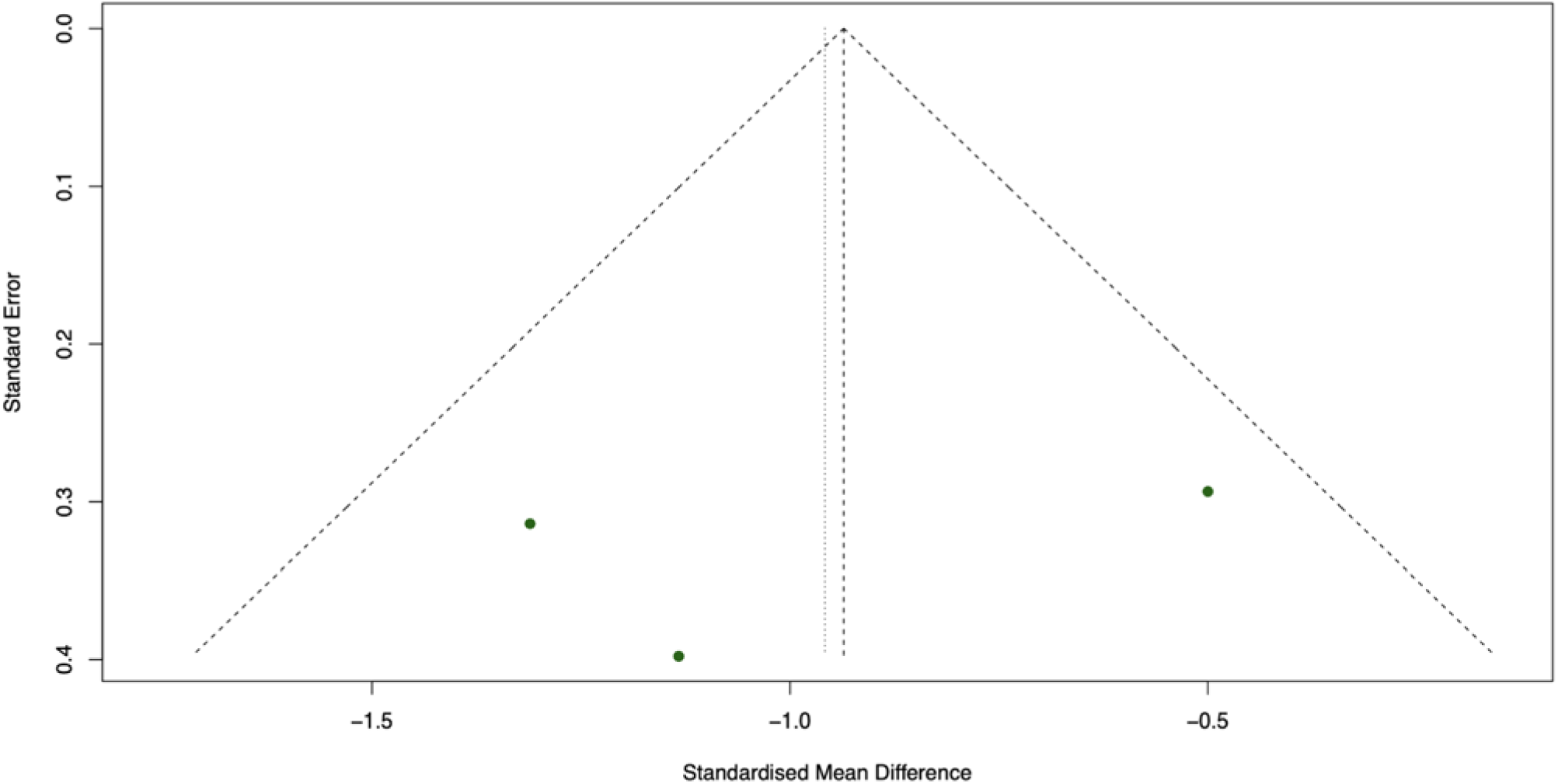
Funnel plot assessing potential publication bias in the alpha diversity meta-analysis.

**Figure 5.**
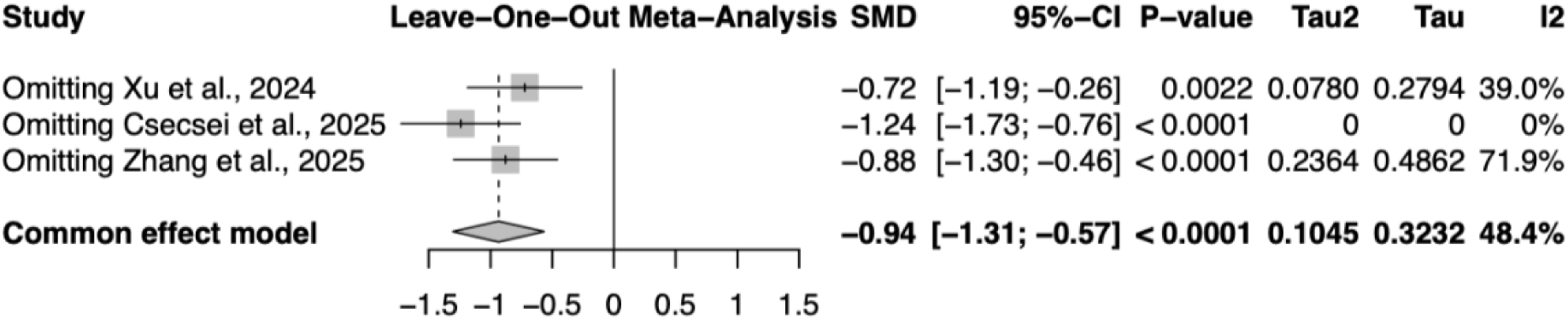
Leave-one-out sensitivity analysis for the alpha diversity meta-analysis.

**Figure 6.**
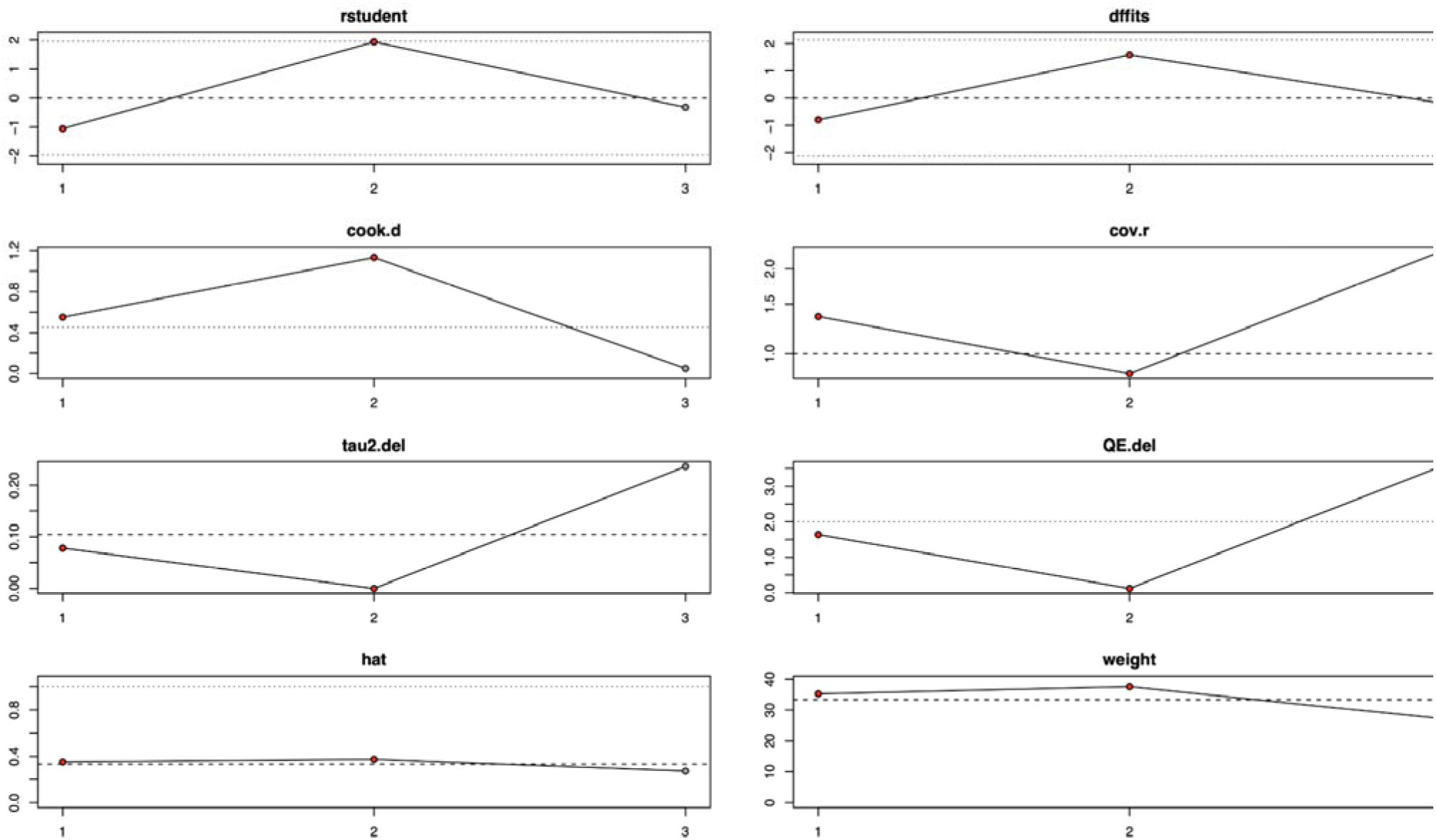
Influence diagnostics for the alpha diversity meta-analysis.

### Risk of bias

Risk of bias assessment using the adapted QUIPS tool showed that most of the 20 included studies had overall low to moderate methodological risk. Study attrition, exposure and outcome measurement, and statistical analysis domains generally scored low risk due to validated sequencing methods, imaging-confirmed aneurysm status, and appropriate regression and MR sensitivity analyses. The main limitation across studies was confounding: although vascular risk factors were commonly adjusted for, key microbiome-related confounders, such as diet, antibiotics, and medication use, were inconsistently controlled. Observational cohorts demonstrated strong measurement quality but variable representativeness and potential selection bias from strict exclusion criteria. Mendelian randomization studies scored low risk for measurement but moderate risk for participation and confounding due to inherent constraints of summary-level genetic data. Overall, risk of bias was acceptable but highlights the need for better control of microbiome-specific confounders in future studies. (fig.2)

### Mendelian Randomization Meta-Analysis of Protective Microbial Taxa

Four microbial taxa consistently reported across independent Mendelian randomization studies were included in pooled analysis: Ruminococcus1, Bilophila, Fusicatenibacter, and Porphyromonadaceae. Meta-analysis demonstrated protective associations between genetically predicted abundance of these taxa and aneurysm-related outcomes. The pooled odds ratio for Ruminococcus1 was 0.49 (95% CI 0.35–0.69). For Bilophila, the pooled odds ratio was 0.68 (95% CI 0.55–0.85). For Fusicatenibacter, the pooled odds ratio was 0.65 (95% CI 0.50–0.85), and for Porphyromonadaceae, the pooled odds ratio was similarly 0.65 (95% CI 0.50–0.85). No statistical heterogeneity was observed between studies for any of the included taxa (I² = 0%; τ² = 0). The absence of heterogeneity indicates highly consistent effect estimates across studies despite differences in study populations and genetic instruments. These results suggest that increased genetically predicted abundance of these microbial taxa may be associated with reduced risk of aneurysm formation or rupture. (fig. 7)

**Figure 7.**
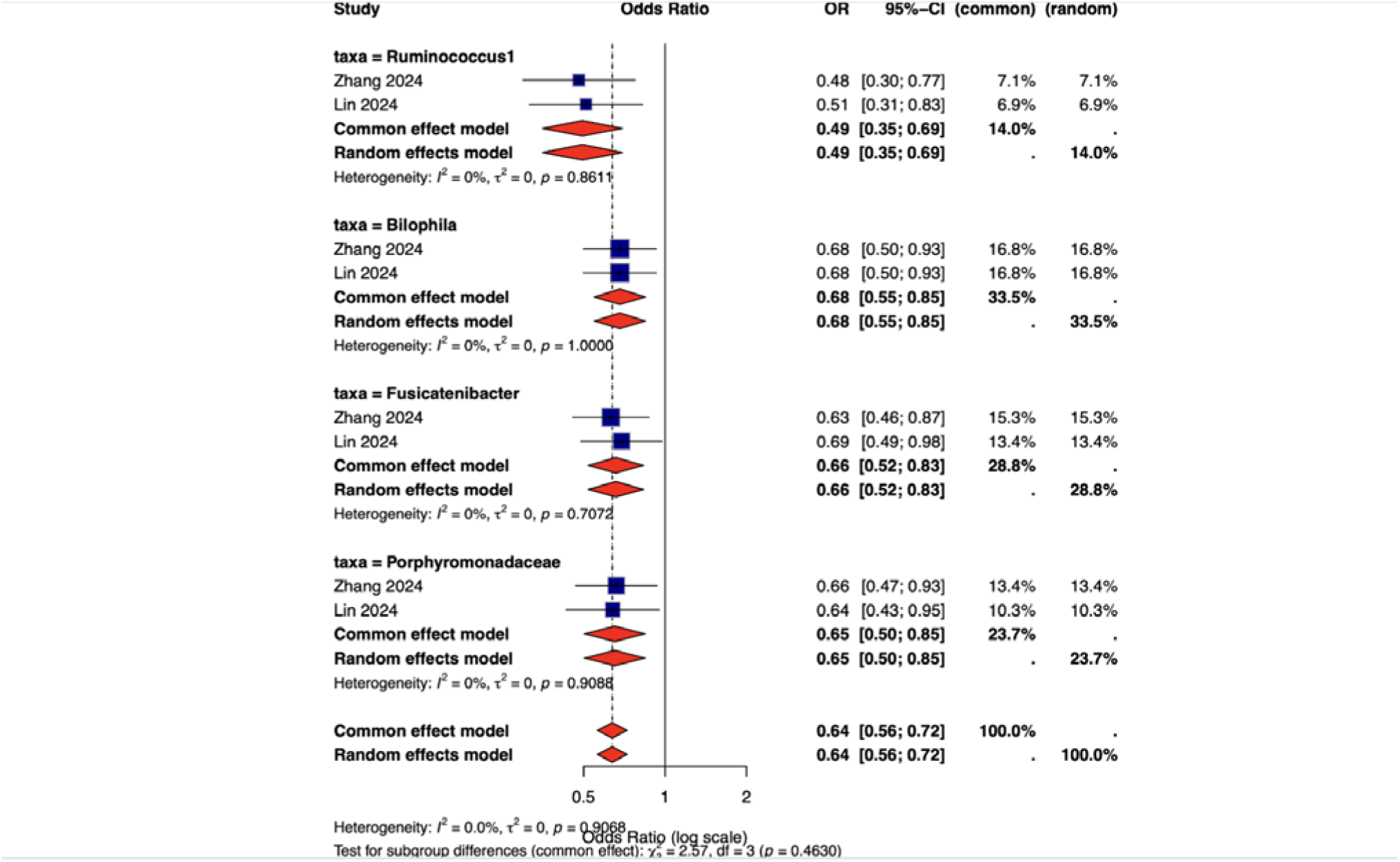
Forest plot summarizing Mendelian randomization meta-analysis results for four protective microbial taxa associated with intracranial aneurysm risk.

## Discussion

### Main Findings

In this study, we performed a comprehensive synthesis of observational microbiome studies, Mendelian randomization analyses, and tissue-based microbial investigations to explore the potential role of the gut microbiome in intracranial aneurysm rupture. Several important findings emerged from our analysis. First, our meta-analysis demonstrated a significant reduction in gut microbiome alpha diversity in patients with ruptured intracranial aneurysms compared with control groups. Pooled analysis of three independent microbiome studies (Xu et al. 2024(17); Csecsei et al. 2025(12); Zhang et al. 2025(18)) including a total of 71 rupture cases and 60 controls showed a significant decrease in microbial diversity in the rupture group, with a pooled standardized mean difference (SMD) of −0.94 (95% CI −1.31 to −0.57) using a random-effects model. Moderate heterogeneity was observed (I² = 48.4%, τ² = 0.1045), suggesting some methodological and population differences across studies but overall consistent direction of effect. Importantly, leave-one-out sensitivity analyses confirmed the robustness of this finding. The pooled effect remained statistically significant after sequential removal of each individual study, with effect estimates ranging from SMD −0.72 to −1.24 and all P values remaining below 0.01. Influence diagnostics further demonstrated that no individual study exerted disproportionate influence on the pooled estimate, supporting the stability of the meta-analytic results. Second, although one of the included microbiome sequencing studies reported no significant differences in alpha diversity between ruptured and unruptured aneurysm patients, significant alterations in microbial community composition were still observed. In the prospective cohort conducted at the University of Pécs (Csecsei et al. 2025)(12), which included 48 patients (24 ruptured and 24 unruptured aneurysms), alpha diversity indices reflecting microbial richness and evenness were comparable between groups. However, beta diversity analyses revealed significant differences in microbial community structure between rupture phenotypes. Specifically, Bray–Curtis dissimilarity analysis demonstrated significant separation between the two groups (p = 0.02), and unweighted UniFrac analysis similarly revealed phylogenetic differences in microbial composition (p = 0.0291). These findings suggest that even in the absence of global diversity loss, shifts in microbial community structure may be associated with aneurysm rupture. Third, differential abundance analyses identified several microbial taxa that differed between ruptured and unruptured aneurysm patients. Genera enriched in the ruptured aneurysm group included Anaerotruncus, Coprobacillus, Sellimonas, Hungatella, and Ruthenibacterium. In contrast, multiple taxa enriched in the unruptured aneurysm group were known short-chain fatty acid (SCFA) producers, including Faecalibacterium, Roseburia, Agathobaculum, members of the Clostridiaceae family, and Brotolimicola. Among these organisms, Faecalibacterium prausnitzii and Agathobaculum butyriciproducens were specifically highlighted in the original study as key butyrate-producing bacteria enriched in patients with unruptured aneurysms. These bacteria are known for their anti-inflammatory properties and their role in maintaining intestinal barrier integrity and vascular homeostasis. Fourth, Mendelian randomization analyses provided additional evidence suggesting potential causal relationships between specific gut microbial taxa and intracranial aneurysm risk. Meta-analysis of taxa identified across independent MR studies (Zhang et al. 2024(25) and Lin et al. 2024(35)) demonstrated consistent protective associations for several bacterial genera. In particular, genetically predicted abundance of Ruminococcus1 was associated with reduced aneurysm risk (pooled OR 0.49, 95% CI 0.35–0.69). Similar protective associations were observed for Bilophila (OR 0.68, 95% CI 0.55–0.85), Fusicatenibacter (OR 0.66, 95% CI 0.52–0.83), and members of the Porphyromonadaceae family (OR 0.65, 95% CI 0.50–0.85). Notably, heterogeneity across these analyses was negligible (I² = 0%), indicating high consistency between the independent MR datasets. Finally, a tissue-based investigation provided additional exploratory evidence supporting a possible link between microbial components and aneurysm pathology. In a cross-sectional study conducted in Brazil, Rabelo et al. (2025)(23) analyzed arterial tissue obtained during microsurgical clipping of intracranial aneurysms and detected bacterial DNA within aneurysm wall samples. Specifically, DNA from Escherichia coli and Fusobacterium nucleatum was identified using polymerase chain reaction. Qualitative analysis suggested a possible association between E. coli DNA detection and aneurysm rupture (OR 4.3, 95% CI 1.01–23.4). However, the statistical analysis reported a P value of 0.9, indicating that the observed association was not statistically significant and should therefore be interpreted with caution. The study’s small sample size and cross-sectional design further limit causal inference. Taken together, these findings suggest that gut microbiome alterations may be associated with intracranial aneurysm rupture through multiple complementary pathways. Reduced microbial diversity, shifts in microbial community composition, depletion of SCFA-producing bacteria, and potential microbial translocation to vascular tissue may collectively contribute to inflammatory and metabolic processes involved in aneurysm instability.

### Comparison With Previous Literature

The relationship between gut microbiota and cerebrovascular diseases has increasingly attracted attention in recent years; however, evidence specifically addressing intracranial aneurysm (IA) and aneurysmal subarachnoid hemorrhage (aSAH) remains limited. Previous systematic reviews have primarily focused on broader cerebrovascular conditions or specific microbial metabolites rather than performing quantitative synthesis of microbiome alterations in aneurysmal disease. A systematic review by Zou et al.(2022)(36) summarized the emerging role of gut microbiota in several cerebrovascular diseases, including ischemic stroke, intracerebral hemorrhage, intracranial aneurysm, and cerebral small vessel disease, emphasizing the importance of the gut–brain axis in cerebrovascular pathophysiology. The authors highlighted several potential mechanisms through which intestinal dysbiosis may influence cerebrovascular pathology, including immune-mediated systemic inflammation, disruption of intestinal barrier integrity, and altered microbial metabolites such as short-chain fatty acids (SCFAs) and trimethylamine-N-oxide (TMAO). Dysbiosis may increase intestinal permeability, promote inflammatory signaling pathways such as Th1/Th17 activation and cytokine release, and contribute to vascular injury and neuroinflammation. However, despite summarizing microbiome alterations across multiple cerebrovascular diseases, the authors acknowledged that evidence specifically related to intracranial aneurysm remains scarce compared with other conditions such as ischemic stroke(36). Importantly, this review did not perform a quantitative meta-analysis and primarily provided a narrative synthesis of available data. Similarly, a systematic review by Klepinowski et al(2023)(13). Evaluated the potential role of the gut microbiome in intracranial aneurysm growth, aneurysmal subarachnoid hemorrhage, and cerebral vasospasm. Although the authors identified several microbial taxa potentially associated with aneurysmal disease, the number of available studies was extremely limited and the overall evidence base remained heterogeneous. For example, reduced abundance of Hungatella hathewayi was suggested as a potentially protective taxon in patients with intracranial aneurysm, while increased Campylobacter species, particularly Campylobacter ureolyticus, were reported in patients with aneurysmal subarachnoid hemorrhage. However, the review included only a small number of studies and relied on narrative synthesis without quantitative pooling of microbiome diversity or taxonomic findings, limiting the ability to draw robust conclusions regarding the role of gut dysbiosis in aneurysm formation or rupture. In addition to microbiome composition, microbial metabolites have also been investigated in cerebrovascular disease. Zhang et al (2023)(37). conducted a systematic review evaluating the association between the gut microbiota–derived metabolite trimethylamine-N-oxide (TMAO) and stroke outcomes. Their analysis included seven observational studies published between 2019 and 2022, most of which focused on acute ischemic stroke, with only one study involving intracerebral hemorrhage. Notably, the review found that no studies had evaluated the relationship between TMAO and outcomes in subarachnoid hemorrhage, highlighting a significant gap in the literature regarding gut microbiota–related metabolic pathways in aneurysmal disease.

The present systematic review and meta-analysis expands upon these earlier studies in several important ways. First, unlike previous reviews that focused broadly on cerebrovascular diseases or specific microbial metabolites, our study specifically investigated the relationship between gut microbiota and intracranial aneurysm and its clinical phenotypes. Second, our review was conducted in accordance with PRISMA guidelines, with a comprehensive search of PubMed, Scopus, Web of Science, and Embase from database inception to April 2026 without language or time restrictions, and a predefined study protocol. Third, while prior reviews relied primarily on narrative synthesis, our study provides quantitative evidence through meta-analysis of microbiome diversity metrics and integrates emerging evidence including Mendelian randomization analyses. By synthesizing data from recent studies published after earlier reviews, our findings provide a more updated and quantitative assessment of the association between gut microbiota alterations and intracranial aneurysm–related outcomes. Taken together, the current evidence suggests that gut microbiota dysbiosis may play an important role in cerebrovascular pathology through inflammatory pathways, intestinal barrier dysfunction, and microbial metabolites. However, compared with other cerebrovascular diseases such as ischemic stroke, the evidence regarding intracranial aneurysm and aneurysmal subarachnoid hemorrhage remains relatively limited. Our study contributes to addressing this gap by systematically integrating available microbiome studies and providing the first quantitative synthesis focused specifically on aneurysmal disease. In addition to extending previous narrative reviews, our study provides quantitative and causal evidence regarding gut microbiota alterations in intracranial aneurysm. While earlier reviews emphasized the potential role of gut dysbiosis in cerebrovascular diseases without reporting pooled diversity measures(13, 36, 37), our meta-analysis demonstrated a significant reduction in gut microbial alpha diversity among patients with intracranial aneurysm. The pooled random-effects analysis of three studies (Xu 2024; Csecsei 2025; Zhang 2025) showed a standardized mean difference of −0.94 (95% CI −1.31 to −0.57) with moderate heterogeneity (I² = 48.4%). Importantly, the direction of effect was consistent across studies, with individual SMD estimates ranging from −0.94 to −1.31. Sensitivity analysis further confirmed the robustness of this finding, as the association remained statistically significant after sequential exclusion of each study (SMD range −0.72 to −1.24; all p < 0.01). These results provide the first quantitative evidence suggesting that reduced microbial diversity may characterize the gut microbial profile of patients with intracranial aneurysm. Our findings also complement and extend previous literature by integrating Mendelian randomization evidence on specific microbial taxa. While earlier reviews mainly summarized observational associations(13, 36), the MR analyses identified several taxa with potential causal relationships with aneurysm risk. In particular, taxa such as Ruminococcus1, Bilophila, Fusicatenibacter, Porphyromonadaceae, and Bifidobacteriales were associated with protective effects, whereas Ruminococcaceae UCG005, Eggerthella, and Coriobacteriaceae were linked to increased susceptibility to aneurysmal disease. Together with the observed reduction in alpha diversity, these findings suggest that aneurysm-related gut dysbiosis may involve both loss of microbial diversity and compositional shifts toward pro-inflammatory taxa. Notably, prior reviews focusing on microbial metabolites, such as the TMAO-centered analysis by Zhang et al (2023)(37) also highlighted the importance of microbiota-derived pathways in cerebrovascular outcomes but reported a lack of evidence specifically addressing aneurysmal subarachnoid hemorrhage. Our study therefore expands the existing literature by integrating diversity metrics and genetic evidence to better characterize the gut microbiome–aneurysm relationship.

### Biological Interpretation

The available human evidence suggests that disturbances in gut microbial composition, commonly referred to as dysbiosis, may influence vascular inflammation, immune regulation, and endothelial integrity processes to aneurysm development.The gut microbiome plays a role in regulating host, immune and systemic inflammatory function and response (6). One of the most important mechanisms is the production of short-chain fatty acids (SCFAs), including acetate, propionate, and butyrate, which are generated through the fermentation of dietary fiber by beneficial gut microbes (7). These metabolites have been shown to anti-inflammatory effects, maintain intestinal barrier integrity, and contribute to vascular protection by modulating immune signaling and endothelial function (7,8). A reduction in SCFA-producing bacteria, which is commonly observed in dysbiotic, may therefore promote systemic inflammation and endothelial dysfunction, potentially contributing to vascular wall degeneration and aneurysm susceptibility. Experimental evidence supports this hypothesis. Manipulation or depletion of gut microbiota can influence aneurysm formation and rupture risk, suggesting that microbial communities may actively modulate vascular inflammatory processes (9). These experimental findings indicate that gut microbial composition may influence aneurysm pathophysiology through several interconnected pathways, including immune activation, oxidative stress, and inflammatory signaling within the vascular wall. In addition to inflammatory modulation, the gut microbiome may also influence vascular health through systemic cardiovascular risk pathways. Emerging studies have suggested that microbial composition may affect blood pressure regulation, arterial stiffness, and metabolic pathways that contribute to vascular remodeling (9,10). Because hypertension and vascular wall degeneration are well-established contributors to intracranial aneurysm progression, microbiome-mediated alterations in these pathways may indirectly increase the risk of aneurysm instability and rupture.Taken together, these findings support a conceptual framework in which gut microbial dysbiosis may contribute to intracranial aneurysm vulnerability through multiple biological pathways, including reduced production of anti-inflammatory microbial metabolites, increased systemic inflammation, and microbiome-mediated alterations in vascular risk factors.

### Clinical Implications

Gut microbiome profiles could serve as a potential biomarker for aneurysm risk stratification. As highlighted in previous literature, alterations in the gut microbiome may reflect early pathophysiological processes within the gut–brain axis, raising the possibility that microbiome-based signatures could contribute to future diagnostic approaches for cerebrovascular diseases. Furthermore, the identification of specific microbial taxa with protective associations may provide insights into potential therapeutic targets. Previous evidence suggests that modulation of the gut microbiota, through dietary interventions, probiotics, or fecal microbiota transplantation, may influence inflammatory pathways and vascular function(36). Although such strategies remain experimental in the context of intracranial aneurysms, targeting gut dysbiosis may represent a promising avenue for future preventive or adjunctive therapies aimed at stabilizing aneurysms or reducing the risk of rupture. In addition, the microbiome may represent a modifiable risk factor in cerebrovascular disease. Lifestyle factors such as diet, antibiotic exposure, and metabolic conditions effect on gut microbial composition, suggesting that interventions targeting these factors could potentially modify aneurysm-related risk pathways. The gut microbiome may also provide insight into broader systemic mechanisms underlying aneurysm instability. As previous studies have suggested, microbial communities may influence vascular risk factors such as blood pressure regulation and inflammatory activity (9, 10, 38). Understanding these interactions could help clinicians better characterize the systemic biological environment associated with aneurysm progression and rupture. However, the current evidence remains limited, and the clinical translation of microbiome-based strategies will require large prospective studies to clarify causality and determine whether microbiome modulation can meaningfully influence aneurysm progression or rupture risk.

### Future Directions

Future investigations should aim to move beyond current associative evidence and establish mechanistic and causal links between gut microbiome dysbiosis and intracranial aneurysm (IA) formation or rupture. Large, multicenter human cohorts with standardized sampling and sequencing protocols are needed to show changes in gut microbial composition before and after aneurysm rupture. Such longitudinal studies would help distinguish chronic dysbiotic signatures from acute alterations related to hemorrhage, hospitalization, or treatment. Integration of multi-omics approaches will be needed also for mechanistic understanding. Combining metagenomics, metabolomics, and host transcriptomic or genomic data may reveal interactions between microbial metabolites, such as short-chain fatty acids (SCFAs), trimethylamine N-oxide (TMAO), and bile acids, and vascular inflammation, endothelial dysfunction, and extracellular matrix remodeling. These integrated studies can clarify whether SCFA depletion or inflammation-related microbial shifts contribute directly to weakening of aneurysmal walls.Stronger Mendelian randomization (MR) analyses are also required. Future MR studies should use well-powered microbiome genome-wide association datasets, apply genome-wide significance thresholds, perform robust colocalization analyses, and use multivariable MR models accounting for shared metabolic or cardiovascular pathways.Interventional studies represent the next step. Randomized controlled trials assessing dietary fiber enrichment, probiotic or prebiotic administration, and targeted modulation of butyrate-producing taxa could determine microbial restoration influences inflammatory markers or aneurysm progression. Finally, integrating microbial and metabolic biomarkers into existing radiologic and clinical risk-stratification models may refine prediction of aneurysm rupture risk. Collaborative, standardized, and globally inclusive research frameworks are necessary to confirm whether the gut microbiome represents a modifiable causal factor in intracranial aneurysm pathophysiology and to translate these findings into preventive or therapeutic strategies.

### Limitation

Several limitations should be considered when interpreting the results of the meta-analytic components of this study. First, the alpha diversity meta-analysis included only three studies comprising 202 participants. Although the pooled effect size demonstrated a robust and statistically significant reduction in microbial diversity among rupture cases, the small number of eligible studies inherently restricts the precision of the estimates and limits the ability to detect small-study effects or publication bias. Furthermore, methodological heterogeneity was moderate (I² = 48.4%), reflecting differences in study populations, sequencing platforms, sampling time relative to aneurysm rupture, analytical pipelines, and the specific alpha diversity indices used. These factors introduce variability that may not be fully captured through random-effects modeling.

Second, summary statistics from several included studies were not directly comparable due to incomplete reporting of standard deviations, inconsistent reporting of confidence intervals, or lack of raw diversity metrics. This prevented the inclusion of additional potentially relevant studies and limited the comprehensiveness of the quantitative synthesis. Additionally, alpha diversity represents a broad indicator of microbial richness and evenness and may not fully capture ecologically relevant differences in microbial community structure, especially when underlying taxa differ significantly even in the presence of similar diversity levels.

Third, the meta-analysis of protective microbial taxa relied on effect estimates derived from Mendelian randomization studies. Although MR approaches help infer causal directionality, combining multiple MR analyses introduces its own assumptions and constraints. These include differences in genetic instrument selection, underlying ancestry of GWAS datasets, harmonization of microbial quantitative trait loci, and potential violations of MR assumptions such as horizontal pleiotropy. While no statistical heterogeneity was observed across the included MR studies (I² = 0%), the small number of datasets (two per taxon) limits the reliability of heterogeneity tests and prevents assessment of small-instrument bias or directional pleiotropy using advanced MR sensitivity analyses. also, MR studies estimate the effects of genetically predicted microbial abundance rather than directly measured microbiome profiles. These findings should therefore be interpreted as supportive evidence of potential causal pathways rather than direct biological observations. Moreover, many microbiome-related MR analyses use genetic instruments selected at relaxed significance thresholds (e.g., p < 1×10⁻⁵ instead of genome-wide significance at 5×10⁻⁸), which may increase the risk of weak-instrument bias and attenuate causal effect estimates.

Fourth, across both meta-analyses, the number of available human studies remains limited relative to the magnitude of the research question. Many eligible studies were cross-sectional in design, with small sample sizes, variable control group definitions, and limited adjustment for confounding factors such as diet, antibiotic exposure, comorbidities, medication use, and lifestyle factors. These unmeasured or uncontrolled confounders may influence microbiome composition and contribute to residual bias in the observed associations.

Fifth, there were notable differences in patient populations across studies, including geographic location, habitual diet, host genetics, comorbidities, medication exposures, and the clinical state at the time of microbiome sampling (e.g., pre-rupture, post-rupture, acute hospitalization). Timing of stool collection relative to aneurysm rupture varied substantially and was not consistently reported, raising the possibility that acute physiological changes or hospital interventions may have influenced gut microbial composition. These population-level differences likely contributed to observed heterogeneity in taxonomic patterns and diversity indices.

Sixth, the current evidence base for gut microbiome involvement in intracranial aneurysm formation and rupture remains limited in scale and geographic scope. The Introduction section of the manuscript emphasizes that many early human studies were small, conducted in restricted regions, lacked comprehensive control for dietary, lifestyle, and ethnic diversity, and often used short observational windows. These issues reduce external generalizability and highlight the need for broader, standardized research efforts.

Seventh, the limited number of studies prevented subgroup analyses, meta-regression, or assessment of effect modifiers such as aneurysm size, location, patient ethnicity, hypertension status, or microbiome sampling time. These factors could play important roles in shaping observed microbial patterns but could not be explored quantitatively in the current synthesis.

Finally, although this review adhered to PRISMA 2020 guidelines, was prospectively registered in PROSPERO, and involved dual independent review at all screening and extraction stages, residual confounding, unmeasured biases, and the inherent limitations of observational designs cannot be fully eliminated.

## Conclusion

This systematic review and meta-analysis provides a comprehensive synthesis of current human evidence regarding the relationship between gut microbiome alterations and intracranial aneurysm. Across observational studies and Mendelian randomization analyses, the findings may suggest that gut microbial dysbiosis and reduced microbial diversity may be associated with aneurysm rupture and related vascular inflammatory processes. The meta-analysis demonstrated significantly lower alpha diversity in patients with ruptured aneurysms, while MR evidence identified several microbial taxa that may exert protective effects. These findings support the concept of a potential gut–vascular axis in intracranial aneurysm pathophysiology. Microbial metabolites, immune signaling pathways, and inflammation-related mechanisms may contribute to aneurysm wall instability and rupture risk. Importantly, the gut microbiome represents a potentially modifiable factor that could be targeted through dietary interventions, probiotics, or other microbiome-directed therapies. Nevertheless, the current evidence remains limited by heterogeneity in microbiome methodologies, small sample sizes, and a lack of longitudinal human data. Future multicenter prospective studies using standardized microbiome sequencing approaches and integrated multi-omics analyses are needed to confirm causal mechanisms and determine whether microbiome-based biomarkers or therapeutic strategies could improve risk stratification and prevention of aneurysm rupture. Overall, this study highlights the emerging role of the gut microbiome as a promising but still underexplored factor in intracranial aneurysm biology and provides a foundation for future translational research in this field.

## Supporting information

extraction

full result version

full text exclusion file

full texts inclusion file

search strategy

prisma checklist

protocol

rob assesment

title abstract screening

## Data Availability

All data produced in the present study are available upon reasonable request to the authors

## Author Contributions

Farzan Fahim conceptualized and designed the study, developed the study protocol, supervised the research process, prepared the data extraction framework, and critically revised the manuscript. He also served as the corresponding author and guarantor of the study. Mahsa Hemmati contributed to methodological supervision, conflict resolution during study selection, and critical revision of the manuscript. Shahriar Heshmaty and Amin Sharvirani contributed to data extraction and verification of extracted data. Amin Sharvirani and Amin Shahini performed full-text screening and eligibility assessment of potentially relevant studies. Ali Hosseini and Mohammadreza Konarizadeh performed title and abstract screening according to predefined eligibility criteria. Seyyed Mohammad Hosseini Marvast, Faeze Dorisefat, Amirmahdi Mojtahedzadeh, Narges Maham, Abolfazl Omranisarduiyeh, and Farshad Fadaei Juibari contributed to literature review, data organization, and preparation of preliminary study materials. Bahador Malekipour Kashan, Guive Sharifi and Alireza Zali provided senior academic supervision and critical scientific input. Sayeh Oveisi contributed to manuscript editing and final preparation of the article. All authors reviewed and approved the final version of the manuscript.

## Declaration of AI Use

Artificial intelligence (AI) tools (ChatGPT) were used to improve the grammar, clarity, and language of this manuscript. The authors reviewed and approved all content, and take full responsibility for the integrity and accuracy of the work.

## Acknowledgments

The authors would like to express their sincere gratitude to Zahra Bagheri, Ahmad Sharafi, and Sohila Sharafi for their invaluable and unwavering support throughout the development of this work. Their encouragement and assistance played an important role in enabling the completion of this study.

## Appendix and Supplementary Material

- Appendix 1. PRISMA checklist
- Appendix 2. Prospero Protocol
- Appendix 3. Search Strategy
- Appendix 4. Title-Abstract screening excluded studies
- Appendix 5,6. Full text screening Excluded and Included studies
- Appendix 7. Data extraction Sheets
- Appendix 8. Risk of bias
- Appendix 9. Full version of result

## Ethics Approval and Consent to Participate

This study is a systematic review and meta-analysis based exclusively on previously published studies. No primary data were collected and no direct involvement of human participants occurred. Therefore, ethical approval from an Institutional Review Board (IRB) or Ethics Committee was not required.

## Consent to Participate declaration in the manuscript

Not applicable.

## Human Ethics and Consent to Participate declarations

Not applicable.

## Clinical trial number

not applicable.

