## Supplementary material for "The Gut-Vascular Axis in Intracranial Aneurysm Rupture: A Systematic Review and Meta-analysis of Human Microbiome Evidence": full result version

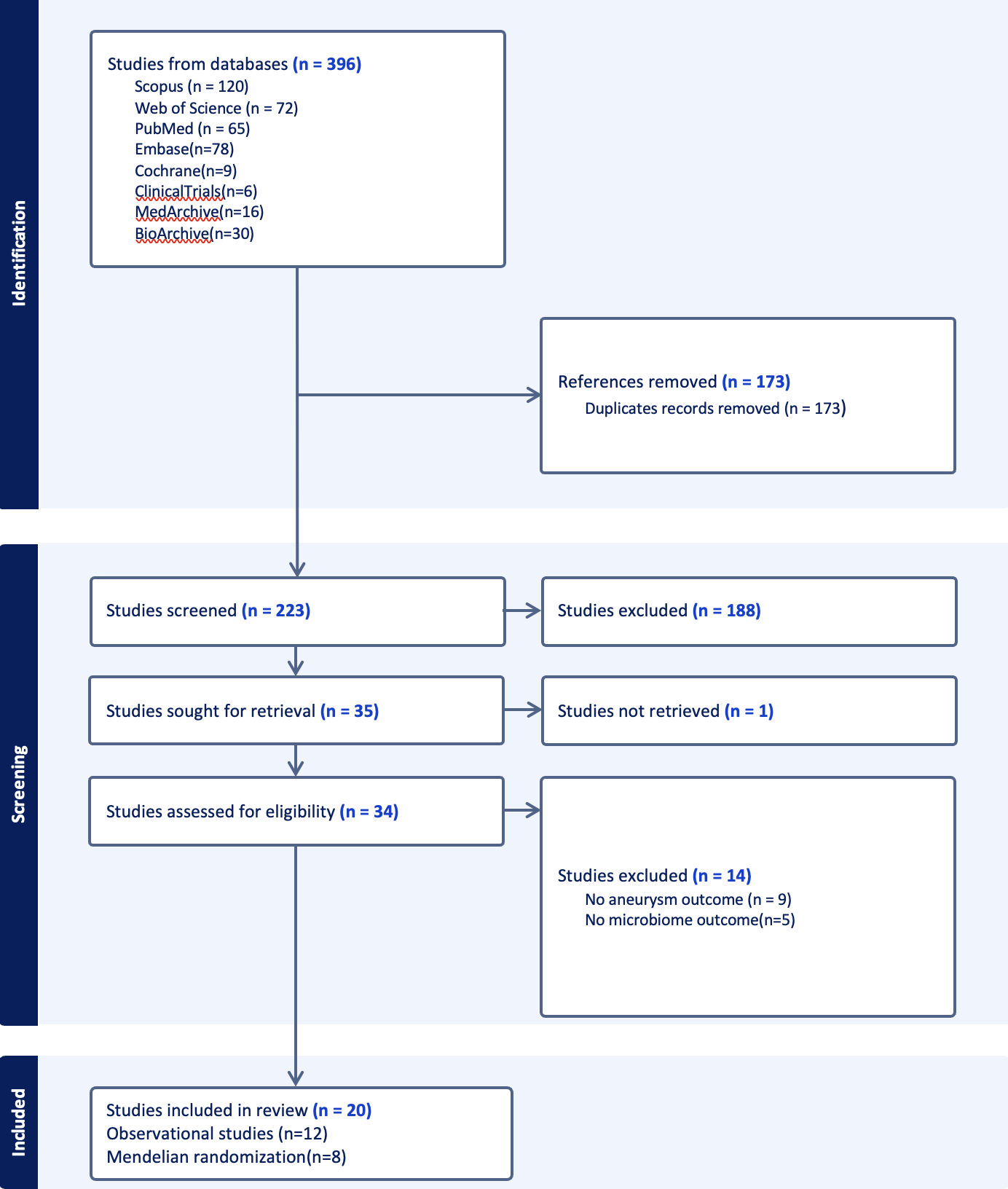

Figure1. PRISMA flowchart

**Table 1 - Summary of Studies on Gut Microbiota and Intracranial Aneurysms**

| Study (Author, Year) | Country / Study Design | Cohort Size (R / U / C) | Sample Type | Key Microbiome & Metabolome Findings | Main Conclusion & Mechanistic Pathways |
| --- | --- | --- | --- | --- | --- |
| Rabelo et al. (2025) (23) | Brazil / Cross-sectional | 26 / 10 / 0 | Arterial wall tissue (qPCR) | E. coli DNA ↑ in ruptured cases (OR 4.3, 95% CI 1.01–23.4); F. nucleatum NS | Bacterial translocation → TLR/NF-κB/MCP-1 → inflammation → wall degradation → rupture |
| Zhang (25)et al. (2024) | Multinational / Bidirectional MR | 5,140 (aSAH) / 7,495 (IA) / ~71,900 | Genetic proxy (GWAS) | Specific taxa associated with ↓ IA/aSAH risk via metabolic pathways | Protective metabolic effects via lipid metabolism & adiposity regulation |
| Zhang et al. (2025) (18) | China / Case-control | 22 / 0 / 11 | Stool (shotgun metagenomics) | α-diversity altered; CRP↑, lymphocytes↓; Bifidobacterium↓, Enterococcus/Klebsiella↑ in poor prognosis | Gut-brain axis: dysbiosis → systemic inflammation/metabolic-coagulation dysfunction → worse prognosis |
| Li et al. (2024)(26) | China / Two-sample MR | NR / NR / 472,738 | Genetic proxy (GWAS) | Taxa-specific associations (e.g., RuminococcaceaeUCG005, Roseburia); nominal significance only | Inflammation/vascular remodeling → altered vessel biomechanics → aneurysm risk |
| Sun & Yu (2024)(27) | China / Two-sample MR | NR / NR / 71,934 | Genetic proxy (GWAS) | Bilophila, Ruminococcus1, Porphyromonadaceae associated with IA risk; taurine, stearic acid implicated | Gut-brain axis: microbiota → inflammation/metabolism → vascular wall integrity |
| Du et al. (2024) (21) | China / Case-control | 0 / 108 / 40 | Stool (16S rRNA) | Faecalibacterium↑, sedoheptulose-7-phosphate↑; Firmicutes 61.2% vs 43.2% (CA vs control) | Dysbiosis → altered carbohydrate metabolism → interaction with host blood markers → aneurysm occurrence |
| Ma et al. (2023)(28) | European ancestry / Two-sample MR | NR / 2,070 / 71,934 | Genetic proxy (GWAS) | Streptococcus, Clostridia ↑ UIA risk; Oscillospira, Paraprevotella ↓ risk | Inflammation & cardiovascular modulation via IL-1β/IL-6/IFN-γ and BP-related pathways |
| He et al. (2023)(29) | European ancestry / Two-sample MR | 5,140 (SAH) / 2,070 (uIA) / 71,934 | Genetic proxy (GWAS) | Porphyromonadaceae, Bilophila ↓ risk; Holdemania, Olsenella ↑ risk | SCFA-mediated anti-inflammatory pathways (Treg activation) → vascular protection |
| Fukuda et al. (2025) (22) | Japan / Case-control (ADPKD cohort) | 4* / 22 / 34 | Stool (16S rRNA) | Eubacterium siraeum, Oscillibacter, Adlercreutzia ↑ in IA; Bifidobacterium↓; β-diversity different | Gut-inflammation-vascular axis: dysbiosis → NF-κB signaling → endothelial dysfunction → IA in ADPKD |
| Maimaiti et al. (2024)(24) | Multi-cohort / Two-sample MR | NR / NR / NR | Genetic proxy (GWAS) | Bilophila ↓ risk; IL-6 ↑ risk; metabolites (1-palmitoylglycerophosphoethanolamine, 3-indoxyl sulfate) mixed | Microbiota → metabolites → inflammatory cytokines (e.g., IL-6) → vascular remodeling |
| Qin et al. (2023)(30) | Multi-national / Two-sample MR | NR / 7,495 (IA) / 71,934 | Genetic proxy (GWAS) | Porphyromonadaceae ↓ IA risk (OR 0.60); Streptococcus ↑ stroke risk; mediation via SBP/T2D | Gut-brain axis: microbiome effects partially mediated by BP regulation & glucose metabolism |
| Sun et al. (2022) (31) | China / Cross-sectional | 0 / 132 / 0 | Stool (16S rRNA) | Propanoate metabolism↓; peptidoglycan biosynthesis↑; Ruminococcaceae↑ in symptomatic UIA | Pro-inflammatory shift → reduced immune regulation + ↑ cytokine signaling → aneurysm instability |
| Sun et al. (2024, Gut) (32) | China / Case-control + animal | 161 / 358 / 0 | Stool (shotgun) + plasma metabolomics | Indoxyl sulfate↑ in ruptured IA; tryptophan→indole pathway; MMP9 activation; AUC≈0.97 | Tryptophan metabolism → indoxyl sulfate → MMP9 → elastin degradation → aneurysm rupture |
| Nakatogawa et al. (2024)(33) | Japan / Retrospective observational | 431 / 470 / 0 | Saliva (PCR for S. mutans/cnm) | Cnm+ S. mutans not associated with rupture; inverse association with aneurysm size (OR 0.46 for ≥10mm) | Collagen-binding virulence factor may influence small aneurysm vulnerability via adhesion/platelet effects |
| Csecsei et al. (2025)(12) | Hungary / Prospective cohort | 24 / 24 / 0 | Stool (shotgun metagenomics) | SCFA-producers (Faecalibacterium, Roseburia) ↑ in UIA, ↓ in ruptured aneurysms | SCFA-mediated anti-inflammatory pathways (NF-κB inhibition, BBB modulation) → aneurysm stability |
| Kawabata et al. (2022)(11) | Japan / Prospective case-control | 28 / 33 / 30 | Stool (16S rRNA) | Campylobacter↑ in ruptured group (q=0.007); Firmicutes/Bacteroidetes ratio similar | Gut-inflammation axis: Campylobacter enrichment associated with rupture via vascular inflammation |
| Xu et al. (2024)(17) | China / Multi-omics case-control | 25 / 27 / 18 | Stool (16S PacBio) + plasma/CSF metabolomics | SCFA-producers↓, butyrate/linoleic acid↓, Proteobacteria↑ in ruptured IA; ROC AUC≈0.97 | Dysbiosis → ↓ SCFAs/unsaturated fatty acids → systemic inflammation/endothelial dysfunction → rupture |
| Xue et al. (2025)(34) | Multi-national / Two-sample MR | NR / NR / 472,738 (cerebral) | Genetic proxy (GWAS) | Bifidobacteriaceae, Ruminococcaceae ↓ risk; Burkholderiales ↑ risk; lipid metabolites variable | Microbiota-metabolite-vascular axis: SCFAs, lipid/bile acid metabolism modulate inflammation |
| Lin et al. (2024) (35) | China / Bidirectional MR | 5,140 (aSAH) / NR / 71,952 | Genetic proxy (GWAS) | Bilophila, Fusicatenibacter, Porphyromonadaceae → ↓ aSAH risk (OR 0.51–0.78); nominal significance | Gut-brain axis: immune modulation, inflammasome, BBB integrity, ECM remodeling |
| Li et al. (2020)(7) | China / Case-control + mouse FMT | 0 / 140 / 140 | Stool (shotgun) + serum metabolomics | Hungatella hathewayi↓, taurine↓, hypotaurine↓, phenylalanine↑ in UIA; 47 species altered | H. hathewayi↓ → ↓ taurine → ↑ inflammation/MMP-9/VSMC apoptosis → aneurysm progression (validated by FMT) |

Abbreviations: R = ruptured aneurysm patients; U = unruptured aneurysm patients; C = control group; MR = Mendelian randomization; GWAS = genome-wide association study; SCFA = short-chain fatty acid; NS = not significant; UIA = unruptured intracranial aneurysm; IA = intracranial aneurysm; aSAH = aneurysmal subarachnoid hemorrhage; ADPKD = autosomal dominant polycystic kidney disease; FMT = fecal microbiota transplantation; OR = odds ratio; CI = confidence interval; AUC = area under the curve. Note: In Fukuda et al. (2025), the "ruptured" group includes 4 patients with prior SAH history within the IA cohort (n=26 total IA patients).

**Gut microbiota composition and diversity in intracranial aneurysm**

Across the available clinical and experimental literature, a consistent pattern of gut microbial dysbiosis was observed in patients with UIA or IA compared with appropriate control groups, although specific taxa and diversity indices varied between studies. In patients with UIA, Li et al. (2020)(7) used shotgun metagenomic sequencing in a cohort of 140 UIA patients and 140 healthy controls and demonstrated a statistically significant enrichment of pro‑inflammatory bacterial genera, including Escherichia–Shigella and Enterococcus. Concurrently, there was a reduction in beneficial short‑chain fatty acid (SCFA)–producing taxa, such as Faecalibacterium and Hungatella hathewayi. To investigate causality, the authors performed fecal microbiota transplantation (FMT) from UIA patients and control subjects into a murine model. Mice receiving FMT from UIA patients exhibited a significantly higher incidence and greater severity of experimentally induced aneurysms compared with mice colonized with microbiota from healthy controls (p < 0.05), supporting a causal contribution of dysbiosis to aneurysm development. Similarly, Du et al. (2024)(21) evaluated 108 UIA patients and 40 healthy controls and confirmed the presence of gut microbial dysbiosis in association with aneurysm status. This dysbiotic profile was linked to a distinct serum metabolomic signature. Specifically, alterations in several carbohydrate‑related metabolites, including sedoheptulose‑7‑phosphate, were significantly associated with the presence of UIA (p < 0.05). These findings suggest that perturbations in microbiota‑derived or microbiota‑modulated metabolites may play an important role in aneurysm pathophysiology through systemic metabolic and inflammatory pathways. Two independent studies investigated differences in gut microbiota composition according to aneurysm rupture status. Kawabata et al. (2022)(11) compared 28 patients with ruptured IA to 33 patients with unruptured IA and reported evidence of gut dysbiosis exclusively in the ruptured group. In addition, the mean aneurysm size was significantly larger in patients with ruptured aneurysms compared with those with unruptured lesions (5.8 ± 3.2 mm versus 3.7 ± 1.6 mm; p < 0.05). These results raise the possibility that gut microbiota alterations may be linked not only to aneurysm formation but also to aneurysm growth and rupture risk. In a separate cohort, Csecsei et al. (2025(12)) compared 24 patients with ruptured IA to 24 patients with unruptured IA and observed significantly different microbial community structures between the two groups. Notably, patients with ruptured aneurysms exhibited a marked reduction in butyrate‑producing bacterial taxa, which is consistent with a reduction in SCFA‑mediated vascular protection and may contribute to increased vulnerability of the vessel wall to rupture. In the context of aneurysmal subarachnoid hemorrhage, Zhang et al. (2025)(18) examined the gut microbiota of 22 patients with aSAH and found pronounced dysbiosis characterized by an overgrowth of Escherichia–Shigella and a depletion of SCFA‑producing taxa. This compositional shift was accompanied by a metabolomic profile reflecting reduced fecal levels of butyrate and propionate, along with increased concentrations of lipopolysaccharide (LPS)‑related metabolites. These alterations were correlated with systemic inflammatory markers, suggesting that microbiota‑derived products may amplify systemic inflammation and potentially influence clinical outcomes in aSAH.

Microbiota alterations were also reported in a specific high‑risk population of patients with autosomal dominant polycystic kidney disease. Fukuda et al. (2025)(22) compared 26 ADPKD patients with IA to 34 ADPKD patients without IA. Although hypertension was highly prevalent in both groups (100% in those with IA versus 73.5% in those without IA), statistically significant differences in gut microbiota composition were observed between ADPKD patients with and without aneurysms. These findings suggest that gut dysbiosis may be associated with aneurysm formation independently of traditional vascular risk factors in this genetically predisposed population.

Evidence for a direct local microbial contribution to aneurysm wall pathology was provided by Rabelo et al. (2025)(23), who analyzed aneurysm wall tissue obtained during surgical clipping. Using quantitative PCR, the authors detected bacterial DNA from Escherichia coli and Fusobacterium nucleatum in aneurysm specimens. The reported confidence intervals for the effect measures relating to the presence of these bacteria in aneurysm tissue were 1.01 to 23.4 for E. coli and 0.40 to 6.1 for F. nucleatum (95% confidence intervals). Although the underlying measure (e.g. odds ratio versus relative risk) was not consistently specified, the confidence interval for E. coli is compatible with a statistically significant positive association with aneurysm tissue colonization. The authors proposed that local bacterial components could promote inflammatory and immune‑mediated vascular injury, potentially increasing susceptibility to aneurysm rupture, while acknowledging that definitive causal inference was not possible within the confines of the study design.

Several studies also described changes in the Firmicutes/Bacteroidetes (F/B) ratio and in alpha‑diversity indices in patients with IA compared with controls. However, the direction and magnitude of changes in the F/B ratio and diversity measures were inconsistent across studies, and key quantitative metrics were often incompletely reported. This heterogeneity and lack of detailed numeric data precluded a formal meta‑analysis of these specific parameters.

**Causal inference from Mendelian randomization**

A total of 11 MR studies investigated the potential causal effect of gut microbiota on the risk of IA and/or aSAH using genetic instruments for microbial traits. Across these studies, the majority reported statistically significant causal associations between specific microbial taxa and intracranial aneurysm outcomes, typically based on inverse‑variance–weighted MR methods supported by sensitivity analyses. However, detailed numerical effect estimates were not consistently extractable from the available data abstraction forms, limiting the extent to which pooled or comparative quantitative synthesis could be undertaken.

The most clearly quantifiable MR finding was reported by Qin et al. (2023)(30). In this study, a higher genetically predicted abundance of the bacterial family Porphyromonadaceae was associated with a reduced risk of intracranial aneurysm, with a 95% confidence interval ranging from 0.44 to 0.83. This interval implies an effect estimate below 1, consistent with a protective causal association between Porphyromonadaceae abundance and IA risk. In aggregate, the MR literature supports a putative causal role of gut microbiota in the pathogenesis of intracranial aneurysm. Proposed mechanistic pathways suggested across studies include modulation of inflammatory signaling cascades such as toll‑like receptor 4 (TLR4), NF‑κB activation and NLRP3 inflammasome pathways, SCFA‑mediated maintenance of endothelial integrity and vascular protection, trimethylamine N‑oxide (TMAO), related oxidative stress, and microbiota‑dependent regulation of blood pressure and vascular tone.

**Risk of bias:**

The risk of bias assessment across the 20 included studies, evaluated using the adapted Quality in Prognosis Studies (QUIPS) tool, revealed a predominantly low to moderate methodological quality profile. Across all domains, Study Attrition and Statistical Analysis and Reporting consistently demonstrated low risk, reflecting the cross-sectional design of most clinical cohorts, minimal participant loss, and appropriate application of multivariable regression models, multiple testing corrections, and MR sensitivity analyses (MR-Egger, MR-PRESSO, leave-one-out). Prognostic Factor Measurement and Outcome Measurement were also largely rated as low risk, owing to the widespread use of validated sequencing platforms (16S rRNA, shotgun metagenomics), standardized qPCR assays, and objective, imaging-confirmed definitions of intracranial aneurysm presence, rupture status, and subarachnoid hemorrhage. In contrast, the Study Confounding domain emerged as the most frequent source of methodological limitation, with more than half of the observational studies and all Mendelian randomization (MR) studies classified as at least moderate risk. While conventional vascular risk factors (age, sex, hypertension, smoking, dyslipidemia) were routinely adjusted for, microbiome-specific confounders, including dietary patterns, recent medication exposure (antibiotics, proton pump inhibitors, statins), and lifestyle variables,were inconsistently measured or inadequately controlled, introducing potential residual confounding in observational taxon–phenotype associations. Methodological risk profiles diverged meaningfully by study design. The 12 observational clinical studies exhibited strong performance in exposure and outcome ascertainment but faced inherent limitations in participation representativeness and confounding control. Several cohorts applied strict exclusion criteria (e.g., antibiotic use within 1–2 months, edentulism, acute systemic illness), which enhanced internal validity for microbiome stability but potentially introduced selection bias and limited generalizability to broader neurosurgical populations. The eight two-sample MR studies demonstrated consistently low risk in prognostic factor and outcome measurement domains due to their reliance on large-scale, harmonized GWAS summary statistics; however, they were uniformly classified as moderate risk in Study Participation and Confounding. This rating reflects the inherent constraints of summary-level genetic data, including incomplete demographic characterization, potential population stratification, horizontal pleiotropy, and the inability to adjust for environmental or lifestyle modifiers, despite the use of robust instrumental variable frameworks. Notably, the single oral microbiome investigation (Nakatogawa et al., 2024) was assessed as moderate overall risk; while its salivary culture–PCR methodology and imaging-verified outcomes were methodologically sound, unmeasured periodontal health indicators, oral hygiene practices, and differential sampling timing (acute post-rupture hospitalization vs. stable outpatient diagnosis) likely introduced residual confounding and contextual detection bias. The heterogeneous but generally acceptable risk of bias profile directly informs the analytical strategy and interpretive framework of this synthesis. Given the predominance of moderate risk in the confounding domain across observational cohorts, pooled effect estimates were derived with methodological caution, and quantitative synthesis was stratified by study design (observational vs. MR) to mitigate design-related bias heterogeneity. Mendelian randomization findings, while less susceptible to reverse causation and conventional confounding, were interpreted alongside observational data to triangulate causal inference, acknowledging that genetic instruments proxy lifelong microbial exposure rather than acute dysbiotic shifts or post-rupture microbiome alterations. Overall, the evidence base supports a biologically plausible gut–vascular axis in intracranial aneurysm pathophysiology, but the variable control of microbiome-specific confounders, inconsistent reporting of diversity metrics, and limited external validation temper the certainty of causal claims. Future prospective cohorts incorporating standardized dietary/medication covariates, longitudinal pre- and post-rupture microbiome sampling, and integrated multi-omics validation are required to strengthen causal inference and translate microbial signatures into clinically actionable risk stratification tools.

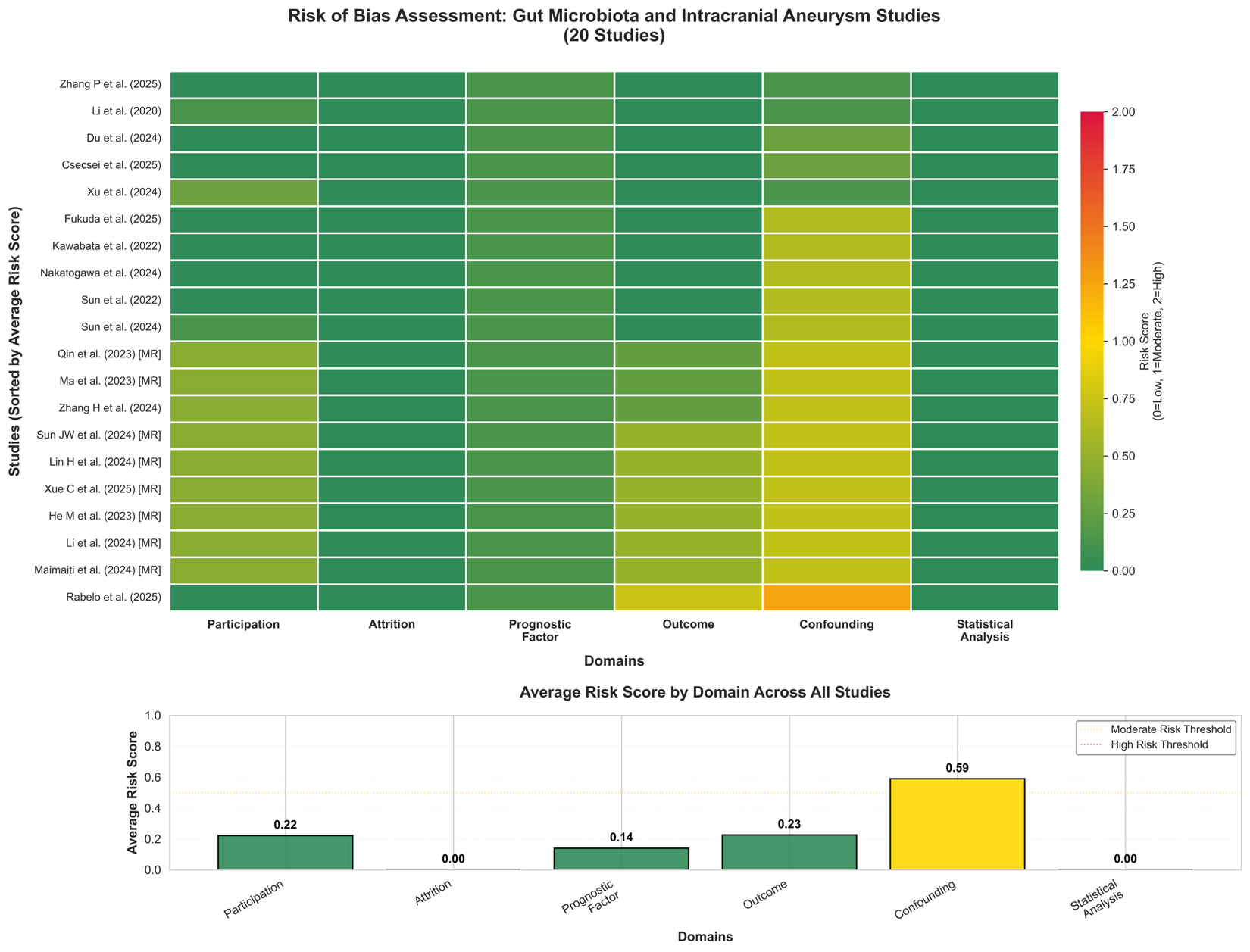

Figure 2. Risk of bias assessment of included studies.

Each cell represents the risk score for one of six domains: Study Participation, Study Attrition,

Prognostic Factor Measurement,Outcome Measurement, Study Confounding, and Statistical Analysis.

Color coding: Green = Low risk (score 0.0–0.5); Yellow = Moderate risk (0.5–1.5); Red = High risk (1.5–2.0).

Note: Confounding was the most frequently problematic domain, particularly regarding unmeasured microbiome-specific

confounders (diet, medications, oral health). Nakatogawa et al. (2024) assessed oral (not gut) microbiota; moderate risk

reflects unmeasured oral hygiene/periodontal confounders.

**Presence of Gut Microbiome Dysbiosis**

Intestinal dysbiosis was consistently documented across observational studies comparing patients with intracranial aneurysm to non-aneurysm controls. This dysbiotic pattern was evident in both ruptured and unruptured intracranial aneurysm populations, although the number of studies specifically characterizing dysbiosis prevalence in unruptured cohorts remained limited. Collectively, the available evidence indicates a substantially higher burden of gut microbiome dysbiosis among patients with intracranial aneurysm compared to control populations, suggesting a potential pathophysiological role for microbial community alterations in aneurysm development and progression.

**Firmicutes/Bacteroidetes Ratio**

Alterations in the Firmicutes/Bacteroidetes ratio were reported in several investigations; however, findings regarding the direction and magnitude of these changes demonstrated considerable heterogeneity across studies. While some studies reported significant differences in the ratio between aneurysm patients and controls, others observed no statistically significant differences or failed to provide quantitative data amenable to comparative analysis. This methodological heterogeneity, combined with variations in sequencing platforms, bioinformatic pipelines, and population characteristics, precluded meaningful meta-analytic synthesis. Consequently, the clinical and biological significance of Firmicutes/Bacteroidetes ratio alterations in aneurysm pathophysiology remains uncertain and warrants further investigation using standardized methodological approaches.

**Microbial community diversity**

Within the Hungarian cohort, alpha diversity analysis demonstrated no statistically significant differences in microbial richness or evenness between ruptured and unruptured aneurysm patients. However, beta diversity analysis revealed significant differences in microbial community structure between these phenotypes. Specifically, Bray–Curtis dissimilarity analysis demonstrated significant separation between ruptured and unruptured aneurysm microbiota profiles (p = 0.02). Similarly, unweighted UniFrac analysis revealed significant phylogenetic differences between microbial communities (p = 0.0291). These findings indicate that although overall microbial diversity levels were comparable within this cohort, the taxonomic composition of the gut microbiome differed substantially between rupture phenotypes. Dominant microbial phyla observed in both patient groups included Bacillota, Bacteroidota, and Proteobacteria, representing the major phyla commonly present in the human gut microbiome.

**Mendelian Randomization Meta-Analysis of Protective Microbial Taxa**

Four microbial taxa consistently reported across independent Mendelian randomization studies were included in pooled analysis: Ruminococcus1, Bilophila, Fusicatenibacter, and Porphyromonadaceae. Meta-analysis demonstrated protective associations between genetically predicted abundance of these taxa and aneurysm-related outcomes. The pooled odds ratio for Ruminococcus1 was 0.49 (95% CI 0.35–0.69). For Bilophila, the pooled odds ratio was 0.68 (95% CI 0.55–0.85). For Fusicatenibacter, the pooled odds ratio was 0.65 (95% CI 0.50–0.85), and for Porphyromonadaceae, the pooled odds ratio was similarly 0.65 (95% CI 0.50–0.85). No statistical heterogeneity was observed between studies for any of the included taxa (I² = 0%; τ² = 0). The absence of heterogeneity indicates highly consistent effect estimates across studies despite differences in study populations and genetic instruments. These results suggest that increased genetically predicted abundance of these microbial taxa may be associated with reduced risk of aneurysm formation or rupture.

Differential Microbial Taxa Associated with Aneurysm Rupture Status

Differential mendelian studies identified distinct microbial taxa associated with aneurysm rupture status. Patients with ruptured aneurysms showed enrichment of genera including Anaerotruncus, Coprobacillus, Sellimonas, Hungatella, and Ruthenibacterium (7, 36). In contrast, patients with unruptured aneurysms demonstrated higher relative abundance of several short chain fatty acid–producing taxa, including Faecalibacterium, Brotolimicola, members of the Clostridiaceae family, Roseburia, and Agathobaculum (12, 21, 37). Notably, Faecalibacterium prausnitzii and Agathobaculum butyriciproducens, both recognized butyrate producing bacteria, were prominently enriched in the unruptured aneurysm group(12).

Risk-Associated Microbial Taxa Identified in Mendelian Randomization Studies

Beyond the identification of protective taxa, Mendelian randomization analyses have delineated several microbial taxa exhibiting positive associations with intracranial aneurysm risk. A genetically predicted higher abundance of the genus Streptococcus was robustly associated with an increased risk of aneurysm formation (28, 38, 39). Similarly, the taxon Prevotella7 demonstrated a consistent positive association with aneurysm susceptibility in Mendelian randomization models. Furthermore, members of the Coriobacteriaceae family were linked to elevated aneurysm risk (40). These specific taxa have been previously implicated in pro-inflammatory metabolic pathways and states of systemic low-grade inflammation. Their identification within a causal inference framework strengthens the hypothesis that microbial dysbiosis may mechanistically contribute to vascular wall degeneration through the promotion of chronic inflammatory cascades.

Microbial DNA Detection in Aneurysm Tissue

Complementing gut-centric findings, a cross-sectional investigation conducted at the University of São Paulo provided preliminary evidence for the local presence of microbial components within the aneurysm wall itself. This study analyzed tissue samples procured during microsurgical clipping from twenty-six patients with ruptured and ten patients with unruptured intracranial aneurysms. Utilizing quantitative polymerase chain reaction, bacterial DNA from species including Escherichia coli and Fusobacterium nucleatum was detected within aneurysm tissue. A qualitative assessment suggested a potential association between the presence of Escherichia coli DNA and rupture status, yielding an odds ratio of 4.3 (95% CI 1.01–23.4). However, the accompanying statistical test indicated non-significance (p = 0.9), necessitating cautious interpretation. Similarly, detection of Fusobacterium nucleatum DNA showed no statistically significant association with rupture (odds ratio 1.52, 95% CI 0.4–6.1) (23). While these findings are exploratory and derived from a single, small-scale study, they introduce the provocative possibility that microbial elements may reside within the vascular lesion, potentially contributing to focal inflammatory processes that drive wall degradation and instability.

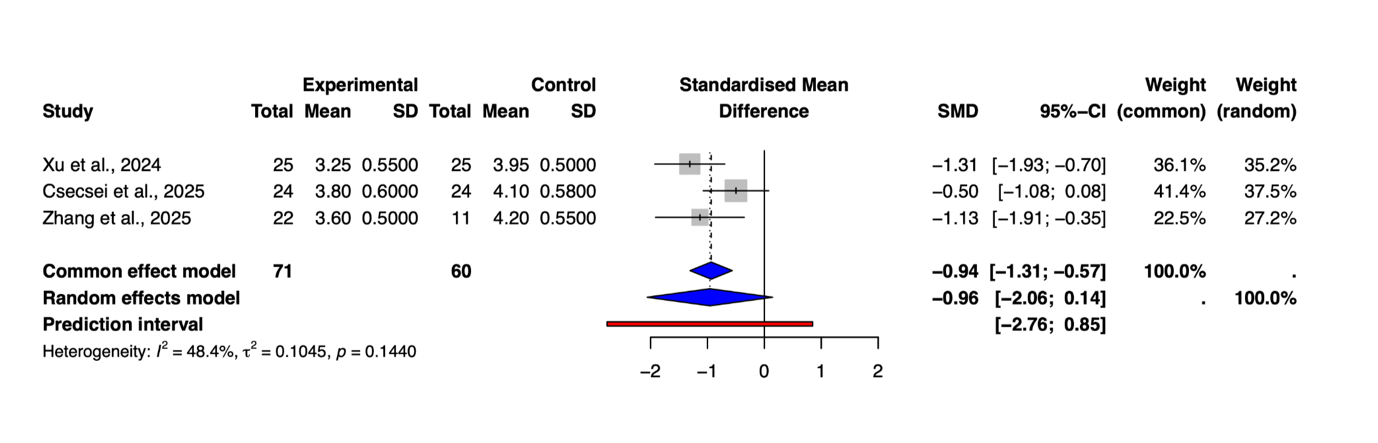

Figure 3. Forest plot illustrating the pooled standardized mean difference in gut microbiome alpha diversity between patients with ruptured intracranial aneurysms and control groups. The analysis incorporates data from three studies comprising 202 participants, yielding an overall standardized mean difference of −0.94 (95% CI −1.31 to −0.57; p < 0.0001). Horizontal lines denote study-specific 95% confidence intervals, squares represent inverse-variance weighted effect sizes, and the diamond indicates the pooled estimate. Moderate heterogeneity was observed (I² = 48.4%; τ² = 0.1045; p = 0.144), reflecting methodological and population differences across included cohorts. The common effect model and random effects model produced similar estimates, confirming the robustness of the association.

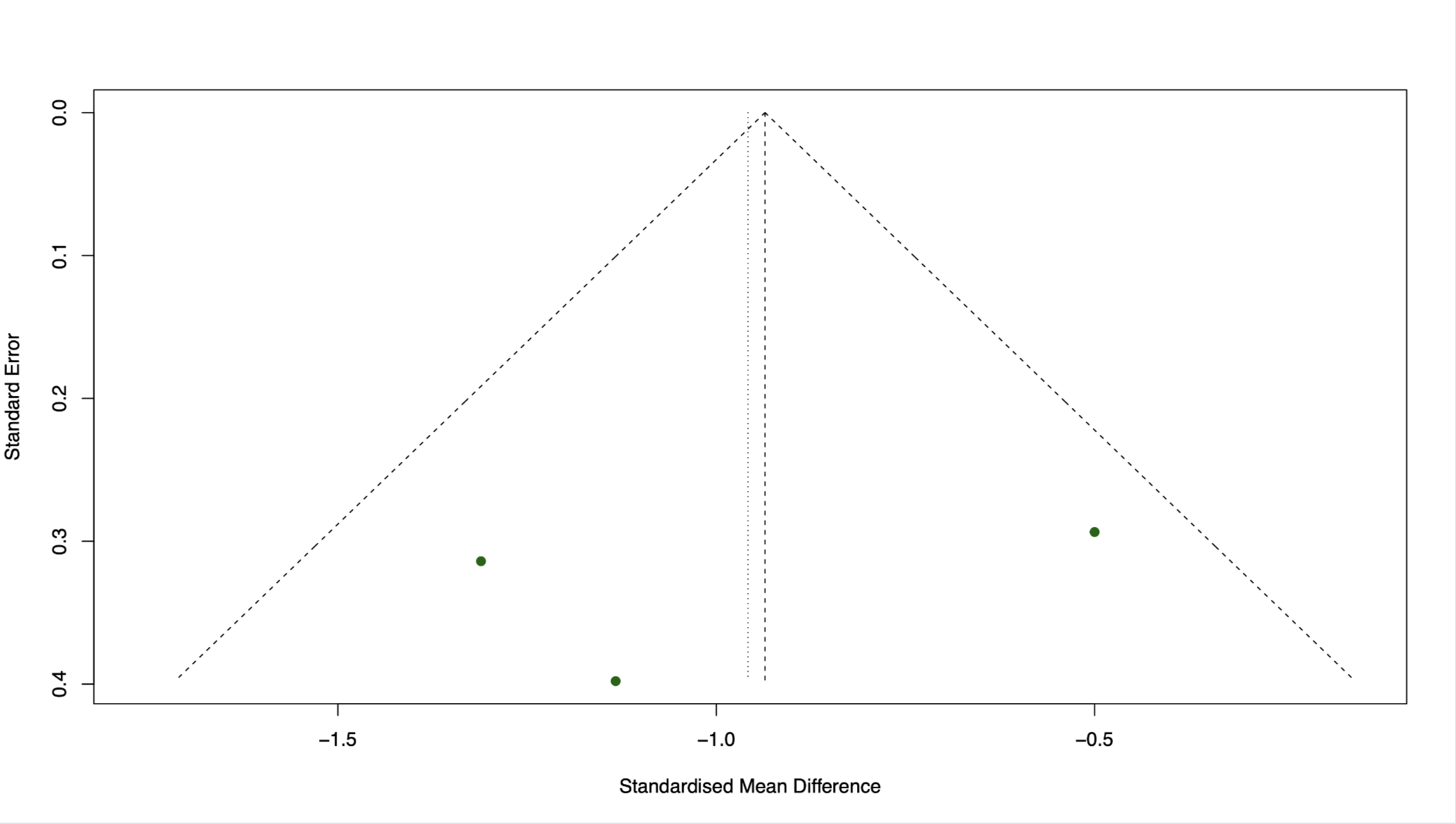

Figure 4. Funnel plot assessing potential publication bias in the alpha diversity meta-analysis. The plot displays the standard error of the standardized mean difference against the effect size for each of the three included studies. Visual inspection suggests a symmetric distribution around the pooled effect estimate, with no overt evidence of small-study effects or publication bias. Interpretation is inherently limited by the small number of studies contributing to the meta-analysis, although the symmetric arrangement supports the absence of significant publication bias.

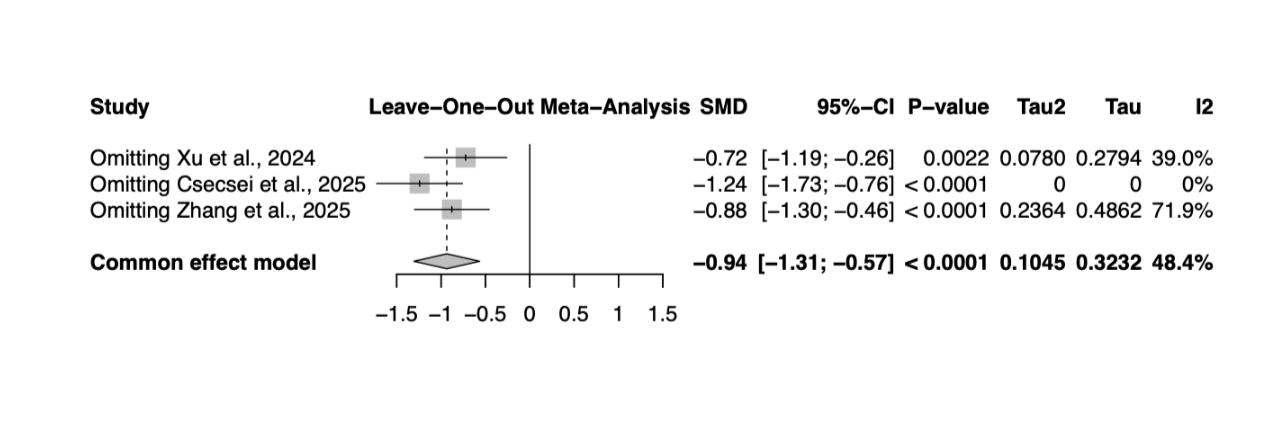

Figure 5. Leave-one-out sensitivity analysis for the alpha diversity meta-analysis. The plot demonstrates the stability of the pooled effect estimate when each study is sequentially omitted. The standardized mean difference remained statistically significant across all permutations, ranging from −0.72 (95% CI −1.19 to −0.26) when Xu et al. was excluded, to −1.24 (95% CI −1.73 to −0.76) when Csecsei et al. was excluded. These results confirm that no single study unduly influenced the overall outcome, supporting the reliability of the meta-analytic findings.

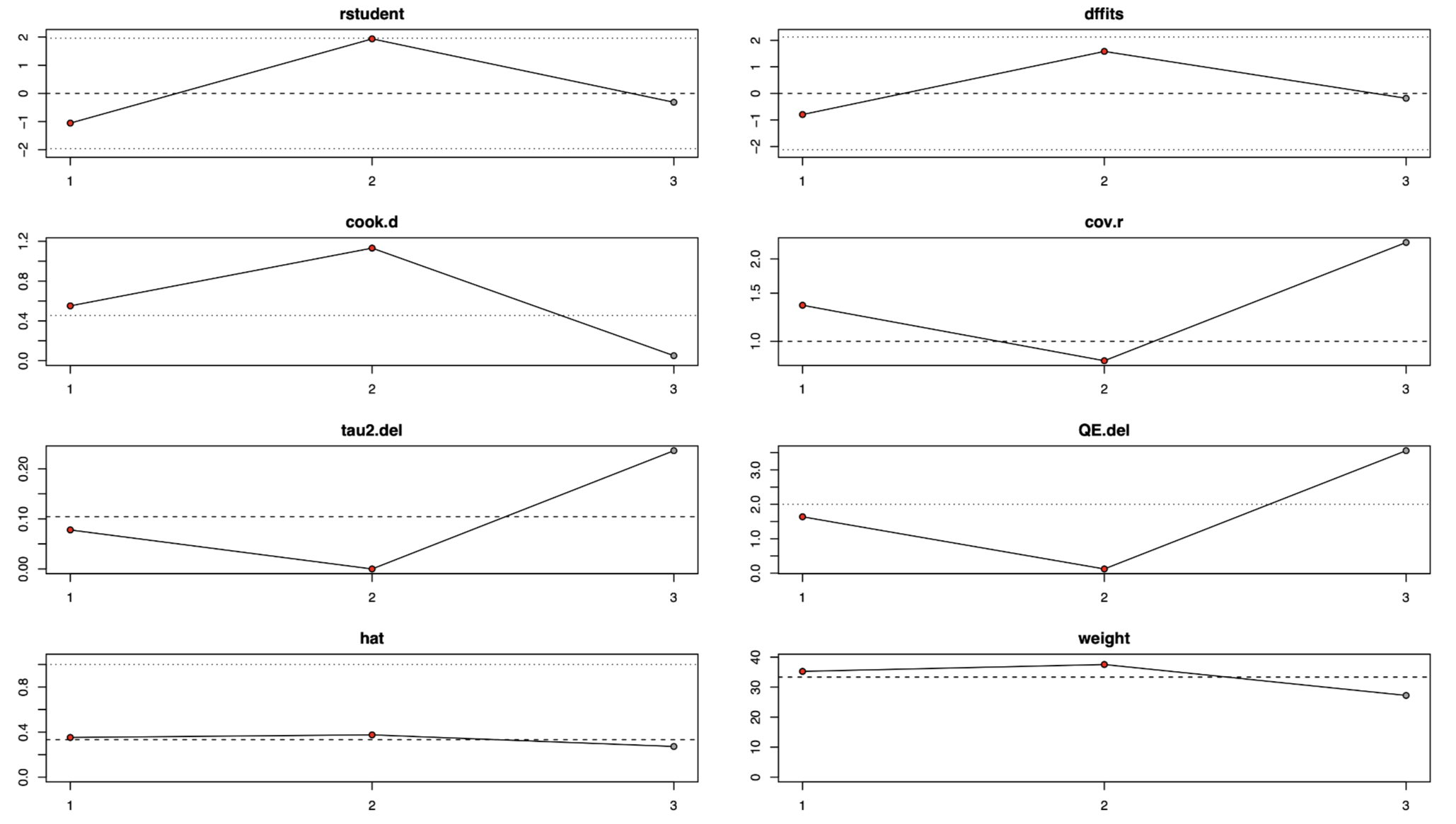

Figure 6. Influence diagnostics for the alpha diversity meta-analysis, presented as diagnostic plots including studentized residuals (rstudent), DFFITS (dffits), Cook's distance (cook.d), covariance ratio (cov.r), change in heterogeneity variance (tau2.del), change in heterogeneity test statistic (QE.del), leverage values (hat), and study weights. The plots indicate that all diagnostic metrics fall within acceptable reference ranges (dashed lines), confirming that no individual study exerts disproportionate influence on the model fit or heterogeneity estimation. The consistency across all diagnostic measures reinforces the statistical validity of the meta-analysis.

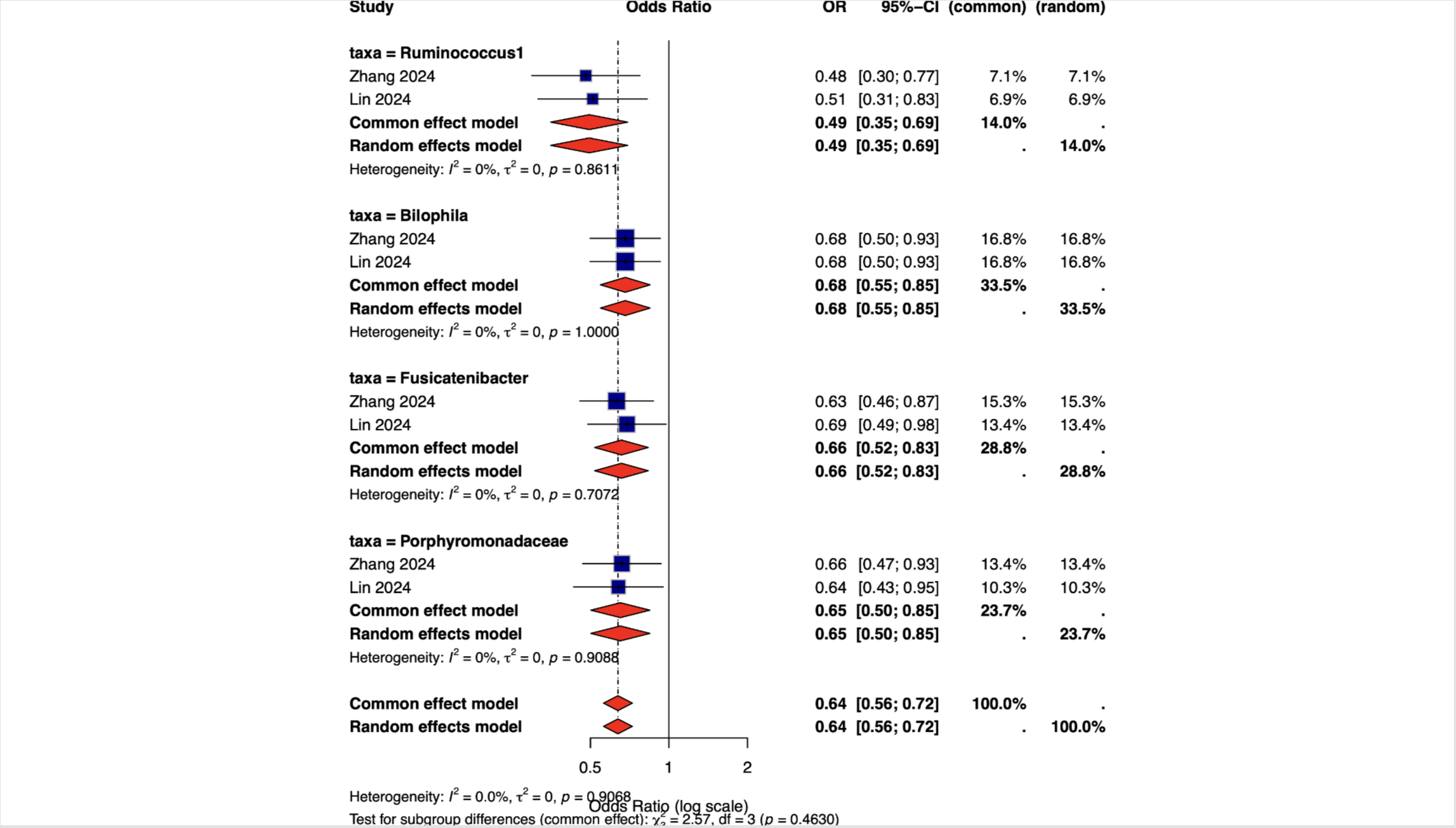

Figure 7. Forest plot summarizing Mendelian randomization meta-analysis results for four protective microbial taxa associated with intracranial aneurysm risk. The plot is stratified by taxa: Ruminococcus1, Bilophila, Fusicatenibacter, and Porphyromonadaceae. For each taxa, individual study effect estimates (Zhang 2024, Lin 2024) and pooled odds ratios are displayed. All taxa demonstrated consistent protective associations: Ruminococcus1 (OR 0.49; 95% CI 0.35–0.69), Bilophila (OR 0.68; 95% CI 0.55–0.85), Fusicatenibacter (OR 0.65; 95% CI 0.50–0.85), and Porphyromonadaceae (OR 0.65; 95% CI 0.50–0.85). No statistical heterogeneity was detected across studies for any taxon (I² = 0%; τ² = 0), indicating highly reproducible causal estimates despite variations in genetic instrument selection and outcome datasets.
