## Supplementary material for "The Gut-Vascular Axis in Intracranial Aneurysm Rupture: A Systematic Review and Meta-analysis of Human Microbiome Evidence": search strategy

**Pubmed**

("Gastrointestinal Microbiome*"[tiab] OR "Microbiome, Gastrointestinal"[tiab] OR "Microflora, GI"[tiab] OR "GI Microflora*"[tiab] OR "Microfloras, GI"[tiab] OR "Microbiome, GI"[tiab] OR "GI Microbiome*"[tiab] OR "Microbiomes, GI"[tiab] OR "Enteric Microbiota*"[tiab] OR "Microbiota, Enteric"[tiab] OR "Microbiotas, Enteric"[tiab] OR "Flora, Enteric Microflora*"[tiab] OR "Enteric Microflora Flora*"[tiab] OR "Floras, Enteric Microflora"[tiab] OR "Microflora Flora, Enteric"[tiab] OR “Microflora Floras, Enteric”[tiab] OR “Gut Microflora*”[tiab] OR “Microflora, Gut” [tiab] OR “Gastrointestinal Microflora” [tiab] OR “Microflora, Gastrointestinal” [tiab] OR “Gastrointestinal Flora*”[tiab] OR “Flora, Gastrointestinal” [tiab] OR “Gut Flora*”[tiab] OR “Flora, Gut” [tiab] OR “Gastrointestinal Microbial Community*”[tiab] OR “Microbial Community, Gastrointestinal” [tiab] OR “Gut Microbiome*” [tiab] OR “Microbiome, Gut” [tiab] OR “Gastrointestinal Microbiota*”[tiab] OR “Microbiota, Gastrointestinal” [tiab] OR “Microflora*” [tiab] OR “Gut Microbiota*”[tiab] OR “Microbiota, Gut” [tiab] OR “Intestinal Microbiome*”[tiab] OR “Microbiome, Intestinal” [tiab] OR “Intestinal Microflora” [tiab] OR “Microflora, Intestinal” [tiab] OR “Intestinal Flora*”[tiab] OR “Flora, Intestinal” [tiab] OR “Intestinal Microbiota*”[tiab] OR “Intestinal Microbiotas*”[tiab] OR “Microbiota, Intestinal” [tiab] OR “Enteric Bacteria” [tiab] OR “Bacteria, Enteric” [tiab] OR “Gastric Microbiome*”[tiab] OR “Microbiome, Gastric” [tiab])

AND

("Aneurysms, Intracranial"[tiab] OR "Intracranial Aneurysms*"[tiab] OR "Aneurysm, Intracranial"[tiab] OR "Cerebral Aneurysm*"[tiab] OR "Aneurysms, Cerebral"[tiab] OR "Aneurysm, Cerebral"[tiab] OR "Brain Aneurysm*"[tiab] OR "Aneurysm, Brain"[tiab] OR "Aneurysms, Brain"[tiab] OR "Giant Intracranial Aneurysm*"[tiab] OR "Aneurysm, Giant Intracranial"[tiab] OR "Aneurysms, Giant Intracranial"[tiab] OR "Intracranial Aneurysm, Giant"[tiab] OR "Intracranial Aneurysms, Giant"[tiab] OR "Mycotic Aneurysm, Intracranial"[tiab] OR "Aneurysm, Intracranial Mycotic"[tiab] OR "Aneurysms, Intracranial Mycotic"[tiab] OR "Intracranial Mycotic Aneurysm*"[tiab] OR "Mycotic Aneurysms, Intracranial"[tiab] OR "Aneurysm, Anterior Cerebral Artery"[tiab] OR "Anterior Cerebral Artery Aneurysm*"[tiab] OR "Aneurysm, Anterior Communicating Artery"[tiab] OR "Anterior Communicating Artery Aneurysm*"[tiab] OR "Aneurysm, Basilar Artery"[tiab] OR "Aneurysms, Basilar Artery"[tiab] OR "Artery Aneurysm, Basilar"[tiab] OR "Artery Aneurysms, Basilar"[tiab] OR "Basilar Artery Aneurysm*"[tiab] OR "Aneurysm, Middle Cerebral Artery"[tiab] OR "Middle Cerebral Artery Aneurysm*"[tiab] OR "Aneurysm, Posterior Cerebral Artery"[tiab] OR "Posterior Cerebral Artery Aneurysm*"[tiab] OR "Berry Aneurysm*"[tiab] OR "Aneurysm, Berry"[tiab] OR "Aneurysms, Berry"[tiab] OR "Aneurysm, Posterior Communicating Artery"[tiab] OR "Posterior Communicating Artery Aneurysm*"[tiab])

NOT
(Systematic[sb] OR meta-analysis[pt] OR systematic[pt] OR systemtaic review[pt] OR “systemtaic review*”[tiab] OR meta-analysis[tiab] OR narrative review[pt] OR “narrative review”[tiab])

**Scopus**

(TITLE-ABS-KEY("Gastrointestinal Microbiome*" OR "Microbiome, Gastrointestinal" OR "Microflora, GI" OR "GI Microflora*" OR "Microfloras, GI" OR "Microbiome, GI" OR "GI Microbiome*" OR "Microbiomes, GI" OR "Enteric Microbiota*" OR "Microbiota, Enteric" OR "Microbiotas, Enteric" OR "Flora, Enteric Microflora*" OR "Enteric Microflora Flora*" OR "Floras, Enteric Microflora" OR "Microflora Flora, Enteric" OR "Microflora Floras, Enteric" OR "Gut Microflora*" OR "Microflora, Gut" OR "Gastrointestinal Microflora" OR "Microflora, Gastrointestinal" OR "Gastrointestinal Flora*" OR "Flora, Gastrointestinal" OR "Gut Flora*" OR "Flora, Gut" OR "Gastrointestinal Microbial Community*" OR "Microbial Community, Gastrointestinal" OR "Gut Microbiome*" OR "Microbiome, Gut" OR "Gastrointestinal Microbiota*" OR "Microbiota, Gastrointestinal" OR "Microflora*" OR "Gut Microbiota*" OR "Microbiota, Gut" OR "Intestinal Microbiome*" OR "Microbiome, Intestinal" OR "Intestinal Microflora" OR "Microflora, Intestinal" OR "Intestinal Flora*" OR "Flora, Intestinal" OR "Intestinal Microbiota*" OR "Intestinal Microbiotas*" OR "Microbiota, Intestinal" OR "Enteric Bacteria" OR "Bacteria, Enteric" OR "Gastric Microbiome*" OR "Microbiome, Gastric"))

AND

(TITLE-ABS-KEY("Aneurysms, Intracranial" OR "Intracranial Aneurysms*" OR "Aneurysm, Intracranial" OR "Cerebral Aneurysm*" OR "Aneurysms, Cerebral" OR "Aneurysm, Cerebral" OR "Brain Aneurysm*" OR "Aneurysm, Brain" OR "Aneurysms, Brain" OR "Giant Intracranial Aneurysm*" OR "Aneurysm, Giant Intracranial" OR "Aneurysms, Giant Intracranial" OR "Intracranial Aneurysm, Giant" OR "Intracranial Aneurysms, Giant" OR "Mycotic Aneurysm, Intracranial" OR "Aneurysm, Intracranial Mycotic" OR "Aneurysms, Intracranial Mycotic" OR "Intracranial Mycotic Aneurysm*" OR "Mycotic Aneurysms, Intracranial" OR "Aneurysm, Anterior Cerebral Artery" OR "Anterior Cerebral Artery Aneurysm*" OR "Aneurysm, Anterior Communicating Artery" OR "Anterior Communicating Artery Aneurysm*" OR "Aneurysm, Basilar Artery" OR "Aneurysms, Basilar Artery" OR "Basilar Artery Aneurysm*" OR "Aneurysm, Middle Cerebral Artery" OR "Middle Cerebral Artery Aneurysm*" OR "Aneurysm, Posterior Cerebral Artery" OR "Posterior Cerebral Artery Aneurysm*" OR "Berry Aneurysm*" OR "Aneurysm, Berry" OR "Aneurysms, Berry" OR "Aneurysm, Posterior Communicating Artery" OR "Posterior Communicating Artery Aneurysm*"))

AND NOT

(TITLE-ABS-KEY("systematic review" OR "meta-analysis" OR "systematic" OR "narrative review"))

**Web of science**

TS=("Gastrointestinal Microbiome*" OR "Microbiome, Gastrointestinal" OR "Microflora, GI" OR "GI Microflora*" OR "Microbiome, GI" OR "GI Microbiome*" OR "Enteric Microbiota*" OR "Gut Microbiome*" OR "Gut Microbiota*" OR "Intestinal Microbiome*" OR "Intestinal Microbiota*" OR "Gut Flora*" OR "Gastrointestinal Flora*" OR "Intestinal Flora*" OR "Gastric Microbiome*")

AND

TS=("Cerebral Aneurysm*" OR "Intracranial Aneurysm*" OR "Brain Aneurysm*" OR "Basilar Artery Aneurysm*" OR "Middle Cerebral Artery Aneurysm*" OR "Posterior Communicating Artery Aneurysm*" OR "Berry Aneurysm*")

NOT

TS=("systematic review" OR "meta-analysis" OR "narrative review")


**Embase**

('gastrointestinal microbiome'/exp OR 'gut microbiome':ti,ab,kw OR 'intestinal microbiome':ti,ab,kw OR 'gut microbiota':ti,ab,kw OR 'intestinal microbiota':ti,ab,kw OR 'enteric microbiota':ti,ab,kw OR 'gut flora':ti,ab,kw OR 'intestinal flora':ti,ab,kw OR 'gastrointestinal flora':ti,ab,kw)

AND

('intracranial aneurysm'/exp OR 'cerebral aneurysm':ti,ab,kw OR 'brain aneurysm':ti,ab,kw OR 'berry aneurysm':ti,ab,kw OR 'basilar artery aneurysm':ti,ab,kw OR 'middle cerebral artery aneurysm':ti,ab,kw OR 'posterior communicating artery aneurysm':ti,ab,kw)

NOT

('systematic review'/exp OR 'meta analysis'/exp OR 'narrative review':ti,ab,kw)

**Cochrane CENTRAL (Trials)**

("Gastrointestinal Microbiome" OR "Gut Microbiome" OR "Intestinal Microbiome" OR "Gut Microbiota" OR "Intestinal Microbiota" OR "Enteric Microbiota" OR "Gut Flora" OR "Intestinal Flora")

AND

("Intracranial Aneurysm" OR "Cerebral Aneurysm" OR "Brain Aneurysm" OR "Basilar Artery Aneurysm" OR "Middle Cerebral Artery Aneurysm" OR "Berry Aneurysm" OR "Posterior Communicating Artery Aneurysm")

NOT

("systematic review" OR "meta-analysis" OR "narrative review")

**Clinical trial.gov**

(gut microbiome OR gut microbiota OR dysbiosis OR intestinal microbiota OR gastrointestinal microbiome OR microbiome OR microbiota) AND (intracranial aneurysm OR cerebral aneurysm OR brain aneurysm OR subarachnoid hemorrhage)

**Biorxiv/Medrxiv**

(gut microbiome OR microbiota OR dysbiosis) AND (intracranial aneurysm OR cerebral aneurysm OR brain aneurysm OR subarachnoid hemorrhage)
