## Supplementary material for "The Gut-Vascular Axis in Intracranial Aneurysm Rupture: A Systematic Review and Meta-analysis of Human Microbiome Evidence": protocol

NIHR | National Institute for Health and Care Research

PROSPERO
International prospective register of systematic reviews

### The Gut–Vascular Axis in Intracranial Aneurysm Rupture: A Systematic Review and Meta-analysis of Human Microbiome Evidence

*Farzan Fahim, amirmhdi mojtahedzadeh*

**Citation**

Farzan Fahim, amirmhdi mojtahedzadeh. The Gut–Vascular Axis in Intracranial Aneurysm Rupture: A Systematic Review and Meta-analysis of Human Microbiome Evidence. PROSPERO 2026 CRD420261360785. Available from https://www.crd.york.ac.uk/PROSPERO/view/CRD420261360785.

#### REVIEW TITLE AND BASIC DETAILS

##### Review title

The Gut–Vascular Axis in Intracranial Aneurysm Rupture: A Systematic Review and Meta-analysis of Human Microbiome Evidence

##### Condition or domain being studied

p: patinets with cerebral aneurysm

I: gut-brain axis inteventions

o: lowering risk of rupture

##### Rationale for the review

Intracranial aneurysm (IA) is a relatively common cerebrovascular condition, affecting approximately 3.2% of adults worldwide, and its rupture is responsible for nearly 85% of spontaneous subarachnoid hemorrhage (SAH), a catastrophic event associated with high mortality, severe neurological disability, and long-term reduction in quality of life. Although several established risk factors such as smoking, hypertension, alcohol consumption, and genetic predisposition have been identified, the mechanisms determining why some aneurysms remain stable while others progress to rupture are still not fully understood. This uncertainty has prompted increasing interest in additional biological and environmental factors that may influence aneurysm formation and rupture.

In recent years, the gut microbiome has emerged as a potential contributor to vascular and cerebrovascular diseases through its effects on immune regulation, systemic inflammation, and metabolic processes. Beneficial gut bacteria metabolize dietary fiber into short-chain fatty acids (SCFAs), which exert anti-inflammatory effects and help maintain vascular integrity. Disruption of this microbial balance (gut dysbiosis) may reduce the production of protective metabolites while increasing harmful inflammatory byproducts, potentially contributing to vascular wall weakening, aneurysm formation, or rupture. Experimental studies in animal models have supported this hypothesis, demonstrating that modification or depletion of gut microbiota can reduce the likelihood of aneurysm formation and rupture.

##### Review objectives

The objective of this systematic review was to comprehensively evaluate the existing human evidence on the relationship between gut microbiome alterations and intracranial aneurysm formation and rupture. Specifically, this review aimed to assess whether gut microbial dysbiosis, changes in microbial diversity, or variations in microbial composition are associated with the presence of intracranial aneurysms and with differences between ruptured and unruptured aneurysms. In addition, the review sought to summarize reported biological mechanisms, microbial metabolites, and inflammatory pathways that may link gut microbiota to aneurysm pathophysiology. Secondary objectives included examining associations between microbiome characteristics and clinical outcomes such as vasospasm, hydrocephalus, mortality, and functional outcomes following aneurysm rupture.

##### Keywords

Cerebral aneurysm; Gut microbiome; Subarachnoid hemorrhage

##### Country

Iran (Islamic Republic of)

#### ELIGIBILITY CRITERIA

##### Population

*Included*
patients with cerebral aneurysm which is in danger of rupture

##### Intervention(s) or exposure(s)

*Included*
gut microboime changes

##### Comparator(s) or control(s)

This review does not have any comparators

##### Study design

Only nonrandomized study types will be included.

*Included*

cross sectional

cohorts

case controls

##### Context

This systematic review included observational human studies (cohort, case-control, and cross-sectional designs) that evaluated the relationship between gut microbiome alterations and intracranial aneurysm (IA) formation or rupture. Eligible studies were required to involve adult participants with radiologically confirmed IA (CT angiography, MR angiography, or digital subtraction angiography) and to assess gut microbial composition or related metabolites using validated techniques such as 16S rRNA sequencing, shotgun metagenomics, or metabolomic profiling. Studies comparing patients with ruptured aneurysms, unruptured aneurysms, and healthy controls were all eligible. The included studies were conducted in diverse hospital, outpatient, or community settings across Asia, Europe, and North America, reflecting real-world populations. Populations typically consisted of adults aged 40–75 years, with variable distributions of sex, hypertension, smoking, and diabetes, which were extracted as potential confounders. Data items considered included aneurysm size, location, multiplicity, and treatment status, as well as clinical outcomes such as vasospasm, hydrocephalus, rebleeding, mortality, and functional recovery.

Exclusion criteria: animal studies, interventional trials, case reports, case series with fewer than ten participants, conference abstracts, reviews, and editorials.

#### TIMELINE OF THE REVIEW

##### Date of first submission to PROSPERO

05 April 2026

##### Review timeline

Start date: 5 March 2026. End date: 8 May 2026.

##### Date of registration in PROSPERO

05 April 2026

#### AVAILABILITY OF FULL PROTOCOL

##### Availability of full protocol

A full protocol has not been written.

#### SEARCHING AND SCREENING

##### Search for unpublished studies

Both published and unpublished studies will be sought.

##### Main bibliographic databases that will be searched

The main databases to be searched are *Embase.com*, *MEDLINE*, *PubMed* and *Scopus*.

##### Search language restrictions

There are no language restrictions.

##### Search date restrictions

There are no search date restrictions.

##### Other methods of identifying studies

Other studies will be identified by: *searching conference proceedings* and *searching trial or study registers*.

##### Link to search strategy

A full search strategy has been uploaded to PROSPERO. The PDF may be accessed through this link https://www.crd.york.ac.uk/PROSPEROFILES/0807a1e8ba5efec657ed8a6b8b085963.pdf.

##### Selection process

Studies will be screened independently by at least two people (or person/machine combination) with a process to resolve differences.

#### DATA COLLECTION PROCESS

##### Data extraction from published articles and reports

Data will be extracted independently by at least two people (or person/machine combination) with a process to resolve differences.

Authors will not be contacted for further information.

##### Study risk of bias or quality assessment

Risk of bias will be assessed using:

quips

Data will be assessed independently by at least two people (or person/machine combination) with a process to resolve differences.

Additional information will be sought from study investigators if required information is unclear or unavailable in the study publications/reports.

##### Reporting bias assessment

Risk of bias due to missing results will be assessed

##### Certainty assessment

Certainty of findings will not be assessed

#### OUTCOMES TO BE ANALYSED

##### Main outcomes

aneurysm rupture is primary outcome

##### Additional outcomes

alpha diversity changing

protective factors changes

#### PLANNED DATA SYNTHESIS

##### Strategy for data synthesis

A quantitative meta-analysis will be performed when at least two studies report comparable outcome measures and sufficient statistical data. The primary quantitative synthesis will focus on gut microbiome diversity indices, particularly alpha diversity measures comparing patients with ruptured intracranial aneurysms (RIA), unruptured intracranial aneurysms (UIA), and control populations. Effect sizes reported in individual studies (e.g., Shannon index or other alpha diversity metrics) will be extracted along with their standard deviations or confidence intervals. When different diversity metrics are reported across studies, standardized mean differences (SMD) with 95% confidence intervals will be calculated to enable pooled comparison.

A random-effects meta-analysis model will be applied because substantial methodological and clinical heterogeneity is expected across studies, including differences in sequencing methods (e.g., 16S rRNA sequencing or shotgun metagenomics), microbiome sampling strategies (stool, intestinal samples, or tissue), population characteristics, and geographic regions. Between-study heterogeneity will be assessed using the $I^2$ statistic and Cochran's Q test. $I^2$ values of approximately 25%, 50%, and 75% will be interpreted as low, moderate, and high heterogeneity, respectively.

Where sufficient data are available, subgroup analyses will be conducted to explore potential sources of heterogeneity. Planned subgroup analyses include comparisons between ruptured versus unruptured aneurysms, different microbiome sequencing methods, and population characteristics such as geographic region or clinical setting. Sensitivity analyses will also be performed by sequentially removing individual studies to evaluate the robustness of pooled estimates and to assess the influence of single studies on the overall effect size.

For studies reporting associations between specific microbial taxa and aneurysm risk, effect estimates such as odds ratios (ORs) with corresponding 95% confidence intervals will be extracted. These estimates will be summarized narratively and quantitatively when sufficient comparable data are available. Mendelian randomization studies examining genetically predicted gut microbiota traits and intracranial aneurysm outcomes will be analyzed separately from observational microbiome profiling studies due to their different methodological frameworks.

Statistical heterogeneity and consistency of results will be carefully evaluated, and where pooling is not appropriate due to substantial heterogeneity or insufficient comparable data, findings will be synthesized narratively. Publication bias will be assessed visually using funnel plots when at least ten studies are available for a given outcome and statistically using Egger's regression test. All statistical analyses will be conducted using standard meta-analysis software packages.

#### CURRENT REVIEW STAGE

##### Stage of the review at this submission

| Review stage | Started | Completed |
|---|---|---|
| Pilot work | ✓ | |
| Formal searching/study identification | ✓ | |
| Screening search results against inclusion criteria | ✓ | |
| Data extraction or receipt of IPD | | |
| Risk of bias/quality assessment | | |
| Data synthesis | | |

##### Review status

The review is currently planned or ongoing.

##### Publication of review results

Results of the review will be published.

#### REVIEW AFFILIATION, FUNDING AND PEER REVIEW

##### Review team members

**Dr Farzan Fahim** (review guarantor and contact) ORCID: 0000-0003-0591-674X. Shohada-E-Tajrish Hospital. Iran.

No conflict of interest declared.

**Dr amirmhdi mojtahedzadeh**. shohada-e-tajrish hospital. Iran.

No conflict of interest declared.

##### Named contact

##### Review affiliation

shohada-e-tajrish hospital, shahid beheshti university of medical science

##### Funding source

Review has no funding and no agreed support from an academic institution and is done in authors' own time.

##### Peer review

There has been no peer review of this planned review.

#### ADDITIONAL INFORMATION

##### Review conflict of interest

Declared individual interests are recorded under team member details.. No additional interests are recorded for this review.

##### Medical Subject Headings

Brain-Gut Axis; Classification; Dysbiosis; Gastrointestinal Microbiome; Humans; Intracranial Aneurysm

#### SIMILAR REVIEWS

##### Check for similar records already in PROSPERO

*PROSPERO identified a number of existing PROSPERO records that were similar to this one (last check made on 5 April 2026). These are shown below along with the reasons given by that the review team for the reviews being different and/or proceeding.*

- Influence of the Gut Microbiome on Cytochrome P450–Mediated Drug Metabolism in Humans: A Systematic Review [published 14 March 2026] [CRD420261339902]. The review was judged **not to be similar**
- Cancer and the Gut Microbiome: A Systematic Review of Differences Across Cancer Types [published 28 December 2025] [CRD420251275136]. The review was judged **not to be similar**
- Microbiome Connections in Caries Formation: A Systematic Review of Literature on Oral and Gut Flora Interactions [published 2 July 2025] [CRD420250478396]. The review was judged **not to be similar**

##### PROSPERO version history

- Version 1.0, published 05 Apr 2026

##### Disclaimer

The content of this record displays the information provided by the review team. PROSPERO does not peer review registration records or endorse their content.

PROSPERO accepts and posts the information provided in good faith; responsibility for record content rests with the review team. The guarantor for this record has affirmed that the information provided is truthful and that they understand that deliberate provision of inaccurate information may be construed as scientific misconduct.

PROSPERO does not accept any liability for the content provided in this record or for its use. Readers use the information provided in this record at their own risk.

Any enquiries about the record should be referred to the named review contact
